# Blood lead and other metals among pregnant women in Bangladesh: national prevalence, geographic variation and household child exposure

**DOI:** 10.64898/2026.10.12.26365291

**Authors:** Mohammad Khalid Hasan, Fahima Sharmin Hossain, Md Khairul Islam, Ashika Sultana, Md Ashifur Rahman, Saad Mohammad Quaish, Mohiuddin Sharif, Tanzin Naher, S. M. Ashraf Ul Hoque

## Abstract

**Background:** Lead crosses the placenta and harms the developing brain, and no safe level is known. Bangladesh lacks national estimates of lead exposure in pregnancy.

**Objectives:** We estimated blood lead in pregnant women nationally, described where and in whom it was raised, and examined links with child lead and haemoglobin.

**Methods:** In the Bangladesh Multiple Indicator Cluster Survey 2025, 1,932 of 2,711 eligible pregnant women (71%) had valid venous blood results. Survey-weighted modified Poisson models estimated prevalence ratios (PR) for blood lead of 3.5 µg/dL or more, an analytical cut-off. We also studied 540 households with a tested child aged 12 to 59 months.

**Results:** Among tested women, geometric mean blood lead was 2.05 µg/dL (95% CI 1.99 to 2.12). In all, 17.7% (15.7 to 19.9) had 3.5 µg/dL or more and 7.6% (6.1 to 9.3) had 5 µg/dL or more. Greater Dhaka, defined after inspecting district estimates, held 12% of pregnant women but 64% (54 to 73) of those at 5 µg/dL or more. The richest quintile had a crude PR of 3.06 (2.02 to 4.63, P<0.001), attenuated to 1.09 (0.72 to 1.67, P=0.678) after adjustment for place. No clear association was seen with cosmetics, indoor smoking or nearby battery sites. A woman’s lead of 3.5 µg/dL or more was associated with child lead of 5 µg/dL or more (adjusted PR 1.50, 1.16 to 1.96, P=0.002). This cut-off identified only 23% (16 to 30) of them.

**Conclusions:** Raised blood lead in pregnancy varied sharply by place and was highest in greater Dhaka. The crude wealth association was markedly attenuated after adjustment for place. Maternal lead was linked with child lead, but most exposed children lived with women below the cut-off. These findings support source investigation in greater Dhaka and evaluation of targeted lead assessment for women and children.

## Introduction

Lead exposure caused an estimated 5.5 million adult cardiovascular deaths and a loss of 765 million IQ points in young children worldwide in 2019 [1]. No safe blood lead level is known, and the steepest losses in IQ occur at the lowest levels [2]. Exposure before birth is of special concern for the developing brain. Maternal blood lead in the first trimester predicts poorer infant mental development [3]. Lead stored in the mother’s bone is released during pregnancy and crosses the placenta [4]. The fetus can therefore be exposed even when the mother’s current exposure looks low. Clinical guidance recommends follow-up of pregnant women at 5 µg/dL or more [5]. The United States reference value of 3.5 µg/dL describes the upper end of blood lead in young children; it is not a threshold for pregnancy [6].

Lead exposure remains common in low- and middle-income countries, mainly from battery recycling, metal processing, electronic waste and adulterated spices [7]. In rural central Bangladesh, 31% of 430 pregnant women had more than 5 µg/dL [8]. Lead isotope analysis traced much of that exposure to turmeric coloured with lead chromate [9]. A national campaign then cut the share of market turmeric samples with detectable lead from 47% in 2019 to none in 2021 [10]. Other known sources include informal battery recycling [11], industrial air pollution in Dhaka [12,13] and contaminated spices and soil [14]. Pregnant women are also exposed to arsenic in groundwater and to cadmium, which have been linked with smaller birth size in Bangladesh [15,16].

Three gaps remain. First, most studies of pregnant women in Bangladesh come from a few districts and cannot show the national picture or where exposure concentrates. Second, the sources named in local studies have not been tested together in a national sample. Third, it is unclear whether raised lead in a pregnant woman signals exposure of other household members. That question matters because young children share the home environment and are more vulnerable.

The Bangladesh Multiple Indicator Cluster Survey (MICS) 2025 measured lead, arsenic, cadmium and mercury in venous blood of pregnant women for the first time in a national sample [17]. The survey report gave lead and anaemia tables but withheld the other metals because of concerns about detection limits [17]. We aimed to describe blood lead in pregnant women in Bangladesh, and to find where and in which households lead exposure concentrates. We first estimated national lead levels and mapped raised lead across districts, with the other metals as exploratory description. We then identified personal, household and area factors associated with raised blood lead. Finally, we examined whether lead in pregnant women tracked lead in young children of the same household, and maternal haemoglobin.

## Methods

### Study design and data

We did a cross-sectional analysis of the Bangladesh Multiple Indicator Cluster Survey (MICS) 2025. This nationally representative household survey was run by the Bangladesh Bureau of Statistics with UNICEF support [17,18]. It selected 3,149 clusters with probability proportional to size within 131 strata, the urban and rural parts of each district. Twenty households were drawn in each cluster, giving 62,980 households in all 64 districts. Interviews with women aged 15 to 49 years identified those currently pregnant. As this was a secondary analysis, no sample size was calculated for these aims.

### Participants and blood testing

All currently pregnant women in interviewed households were eligible for blood collection, which ran from February to June 2025. Trained phlebotomists collected venous blood after written consent. The International Centre for Diarrhoeal Disease Research, Bangladesh (icddr,b) measured lead, arsenic, cadmium, mercury and haemoglobin in its Dhaka laboratory [17]. The public data and the survey report do not describe the assay, detection limits or quality control. Of 2,711 eligible women, 757 declined consent, 14 consented but gave no sample, and 1,940 gave blood (S1 Fig). Six samples had no valid laboratory result and two had laboratory codes (999 or 9999) rather than values. This left 1,932 women for analysis.

### Outcomes

The primary outcome was blood lead, analysed on the log scale and in categories. No pregnancy-specific reference value exists. We therefore used 3.5 µg/dL as an analytical cut-off, informed by the United States reference value for young children [6]. We also report 5 µg/dL, the level for follow-up in pregnancy [5], and 10 µg/dL. Secondary outcomes were haemoglobin, anaemia and the blood lead of a child in the same household. Anaemia followed the World Health Organization 2024 cut-offs for pregnancy: below 11.0 g/dL in the first and third trimesters and below 10.5 g/dL in the second [21].

Blood arsenic, cadmium and mercury were exploratory outcomes. The data do not state the chemical forms measured, and we assumed total concentrations in whole blood. Blood arsenic reflects recent intake and does not separate inorganic from organic forms. Blood cadmium reflects both recent and longer exposure. For mercury, 5.8 µg/L is a benchmark linked to the United States reference dose for methylmercury [19]; without speciation it serves only as a concentration reference. We report cadmium of 1 µg/L or more only as a description. Values at the lowest reported level were used as reported, without substitution.

### Exposures and covariates

A short form completed at blood collection asked about practices in the previous three months [17]. Items covered use of sindoor and surma, smoking of cigarettes or bidis by anyone inside the home, and eating soil, clay or similar materials. Women reported whether they knew of sites within about 2 km that made, recycled or broke open vehicle batteries. They were also asked about sites that made or recycled jewellery, recycled electronic waste or repaired automobiles. A further item asked whether anyone in regular contact with the woman worked at such a site. Women also reported hours spent outside the home on the previous day. We coded each item as any versus none, except indoor smoking, which had three levels. Covariates were age, education, previous live births, trimester from the reported week of gestation, wealth quintile, urban residence and division.

### Statistical analysis

All estimates used the women’s blood sample weight and accounted for clustering and stratification. Women outside each analysis group were kept in the design through subpopulation estimation. Geometric means and their 95% confidence intervals came from survey means of log concentrations. Confidence intervals for proportions used the logit transformation. We report exact P values from Wald tests. We first reproduced the published report tables for lead and anaemia. All analyses were done in Stata 17, and figures were drawn in R 4.6.1.

#### Levels and geography of blood lead

Weighted percentiles described the upper tail of each metal. We estimated lead by area, division, wealth quintile and other characteristics. We produced district estimates with 95% confidence intervals after merging city corporations into their districts. The district map showed a block of high prevalence around Dhaka. We therefore defined greater Dhaka as Dhaka district, Dhaka North and Dhaka South City Corporations, Gazipur and Narayanganj (MICS district codes 26, 25, 20, 33 and 67). This follows the usual metropolitan boundary, but the grouping is data-derived and exploratory. We estimated the share of all pregnant women, and of those with raised lead, who lived in greater Dhaka, with 95% confidence intervals. To check blood arsenic, we related it to the field arsenic test of the household’s source water. That test was done in two households per cluster, so the comparison used 201 women.

#### Factors associated with raised blood lead

We fitted survey-weighted Poisson models with robust variance to estimate prevalence ratios for blood lead of 3.5 µg/dL or more [20]. Model 1 included all covariates and all exposure items except eating soil, which only 17 women reported. Model 2 replaced division with place in three groups: greater Dhaka, the rest of Dhaka division and other divisions. To see how far place accounted for the wealth gradient, we started from a wealth-only model. We then added urban residence and division, then greater Dhaka, and finally replaced division with district fixed effects. We also fitted linear models of log blood lead and report geometric mean ratios. We fitted Model 1 again among women outside greater Dhaka. Model 1 had 31 terms for 254 women with raised lead. We therefore checked overlap among the site items with correlations and variance inflation factors. We also fitted a simpler model with place, wealth, education, age and trimester.

#### Links with child lead and haemoglobin

MICS 2025 also tested one randomly selected child aged 12 to 59 months in each household and collected soil near the dwelling. We linked each woman to the tested child of her household; the woman was not always the child’s mother. Pair analyses used the woman’s blood weight. Their target population is households with a pregnant woman and a tested child aged 12 to 59 months. We compared women with and without a tested child, and repeated pair models with the child’s blood weight. We report the Spearman correlation and the ratio of child to woman geometric means. A survey-weighted linear model regressed log child lead on log woman lead, adjusted for child age, wealth, urban residence and division. A further model added household soil lead. We estimated the prevalence ratio for child lead of 5 µg/dL or more when the woman had 3.5 µg/dL or more. We repeated these analyses for pairs in which the woman was the child’s mother. To judge maternal lead as a marker of child lead, we estimated weighted sensitivity, specificity and predictive values. The marker was a woman’s lead of 3.5 µg/dL or more, and the target was child lead of 5 µg/dL or more.

We regressed haemoglobin and anaemia on log lead and on lead of 3.5 µg/dL or more. A further model entered all four metals together; its arsenic, cadmium and mercury terms are exploratory. These models adjusted for age, education, previous births, trimester, wealth, area and division.

#### Missing data and sensitivity analyses

Of 2,711 eligible women, 1,932 (71.3%) had valid metal values. In our check, the blood weight equalled the women’s weight divided by the response rate of eligible pregnant women in each stratum, then rescaled. It therefore corrects for differences in response between strata, but not within them. We modelled the probability of a valid test from age, education, previous births, trimester, wealth, area, division and religion, weighted by the women’s design weight (S10 Table). Dividing the women’s weight by this probability gave response weights, which we also trimmed at the 1st and 99th percentiles. This assumes that, within these groups, testing was unrelated to blood lead, which cannot be checked. We repeated the prevalence, greater Dhaka, Model 1, Model 2 and pair estimates with these weights. Other checks used the 5 µg/dL outcome and added religion, cooking fuel, water source and month of collection. We also added household soil lead where a valid soil result existed (534 women), and excluded values at the lowest reported level. Models used complete cases (1,921 women, 254 with blood lead of 3.5 µg/dL or more). We did not adjust for multiple testing, so P values for secondary and sensitivity analyses are exploratory.

#### Ethics

MICS 2025 was approved by the icddr,b Ethical Review Committee (PR-24113) [17]. Participants gave written informed consent. We used anonymised public data obtained from UNICEF with permission.

## Results

### Participants

Of 2,711 eligible pregnant women, 1,940 (71.6%) gave blood and 1,932 had valid metal values (S1 Fig, S1 Table). Valid tests were less common among richer women (54.6% in the richest quintile against 82.2% in the poorest). They were also less common in Dhaka division (61.2%) and among women with higher secondary education (61.1%). After adjustment, the odds of a valid test were 0.38 (0.26 to 0.56) in the richest quintile and 0.51 (0.36 to 0.74) with higher secondary education (S10 Table). Our estimates matched the published report to within 0.1 percentage points for every lead category, for the urban, rural and wealth gradients, and for anaemia (S2 Table). Women had a mean age of 24.3 years and a mean of 20 weeks’ gestation (Table 1). Over two fifths (42.0%) lived in a home where someone smoked almost daily. Use of surma was reported by 29.5% and sindoor by 9.2%.

**Table 1.**
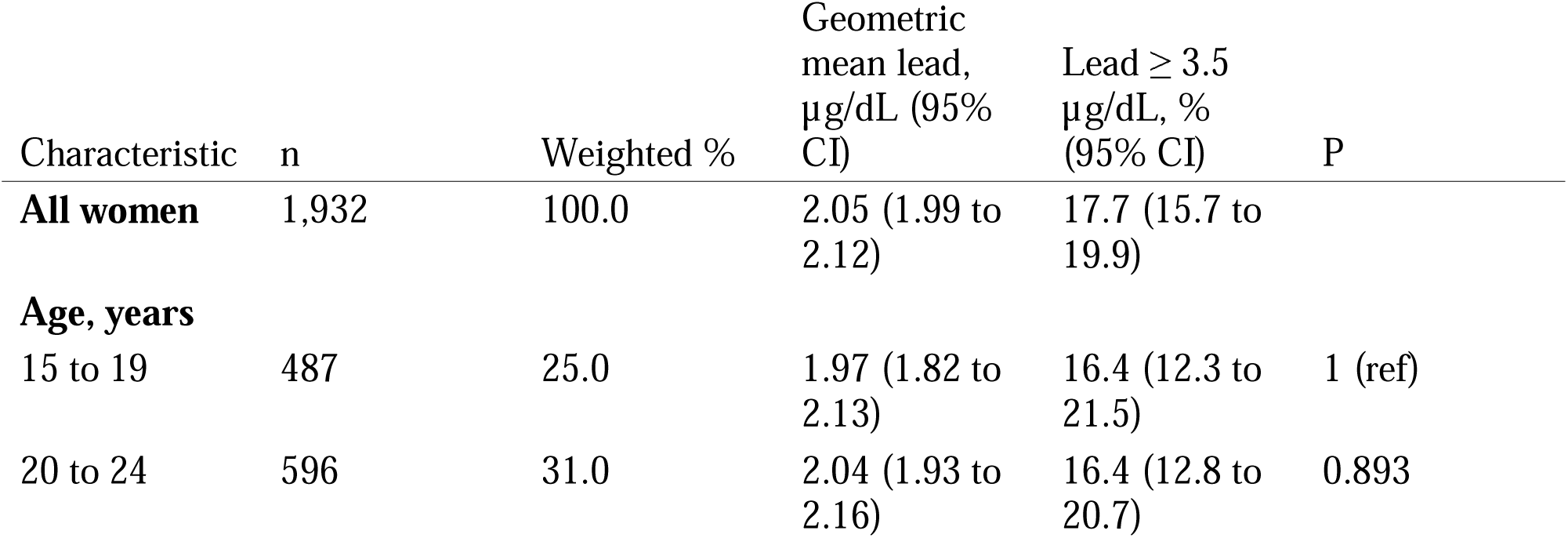
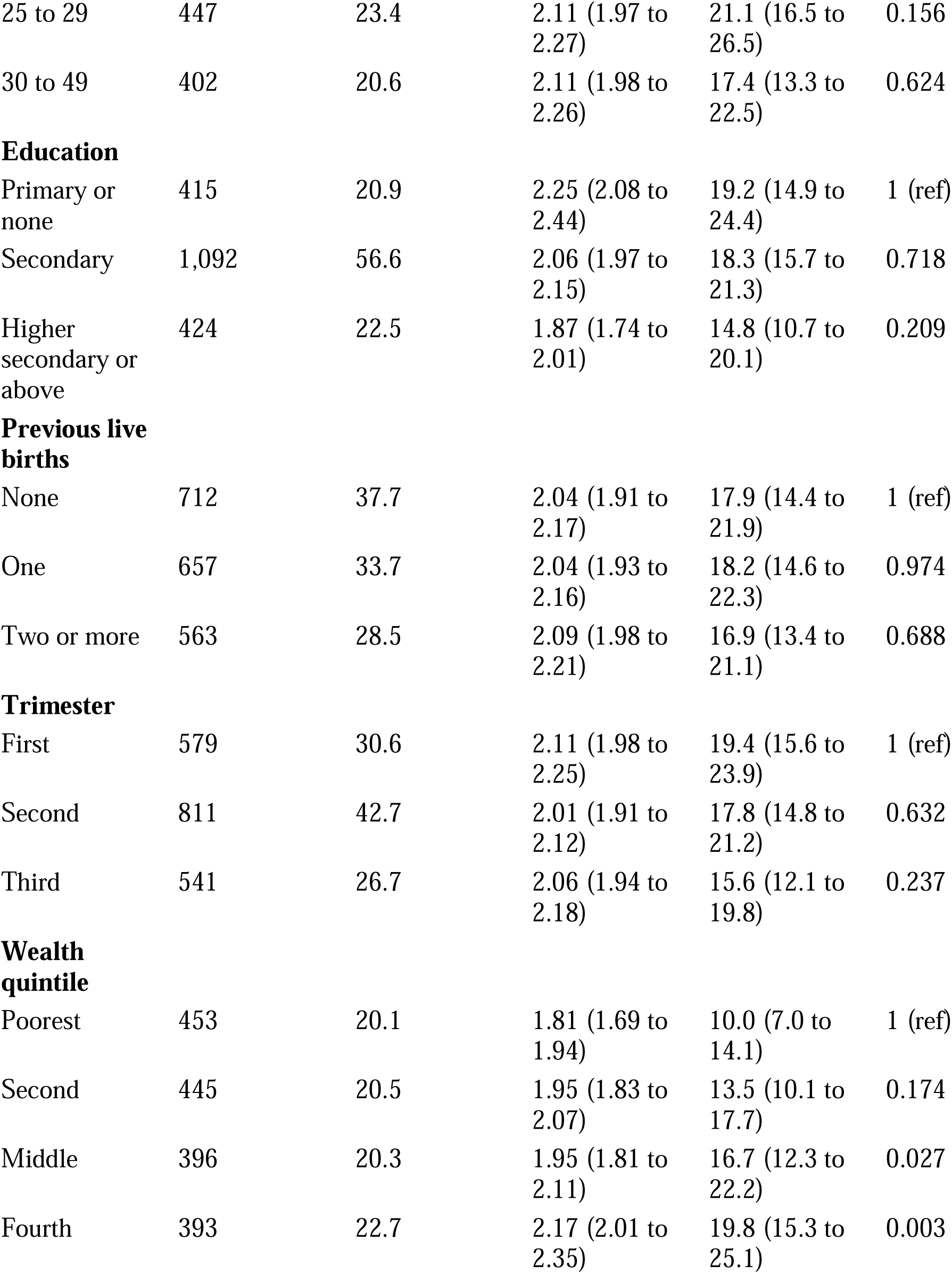
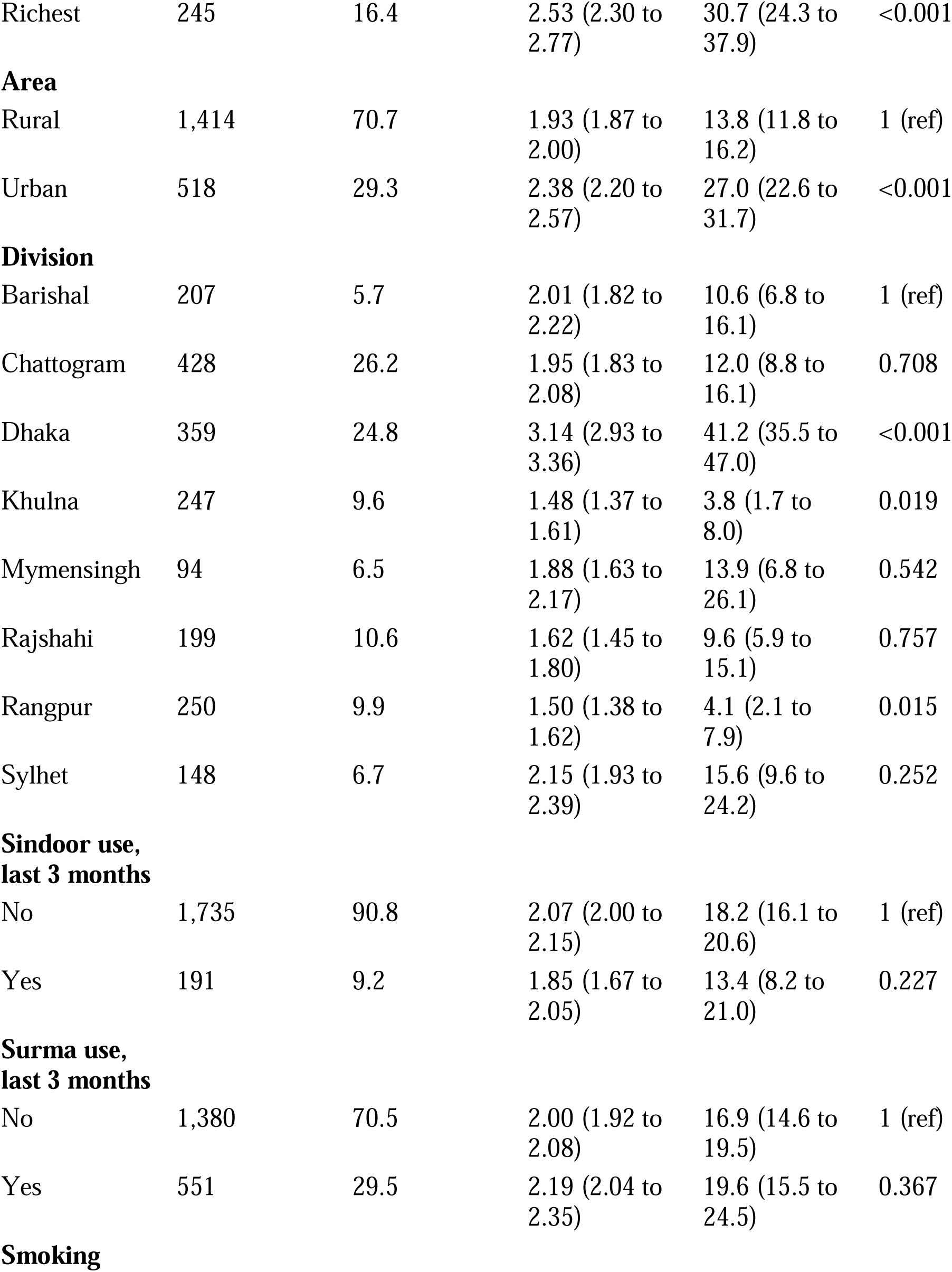
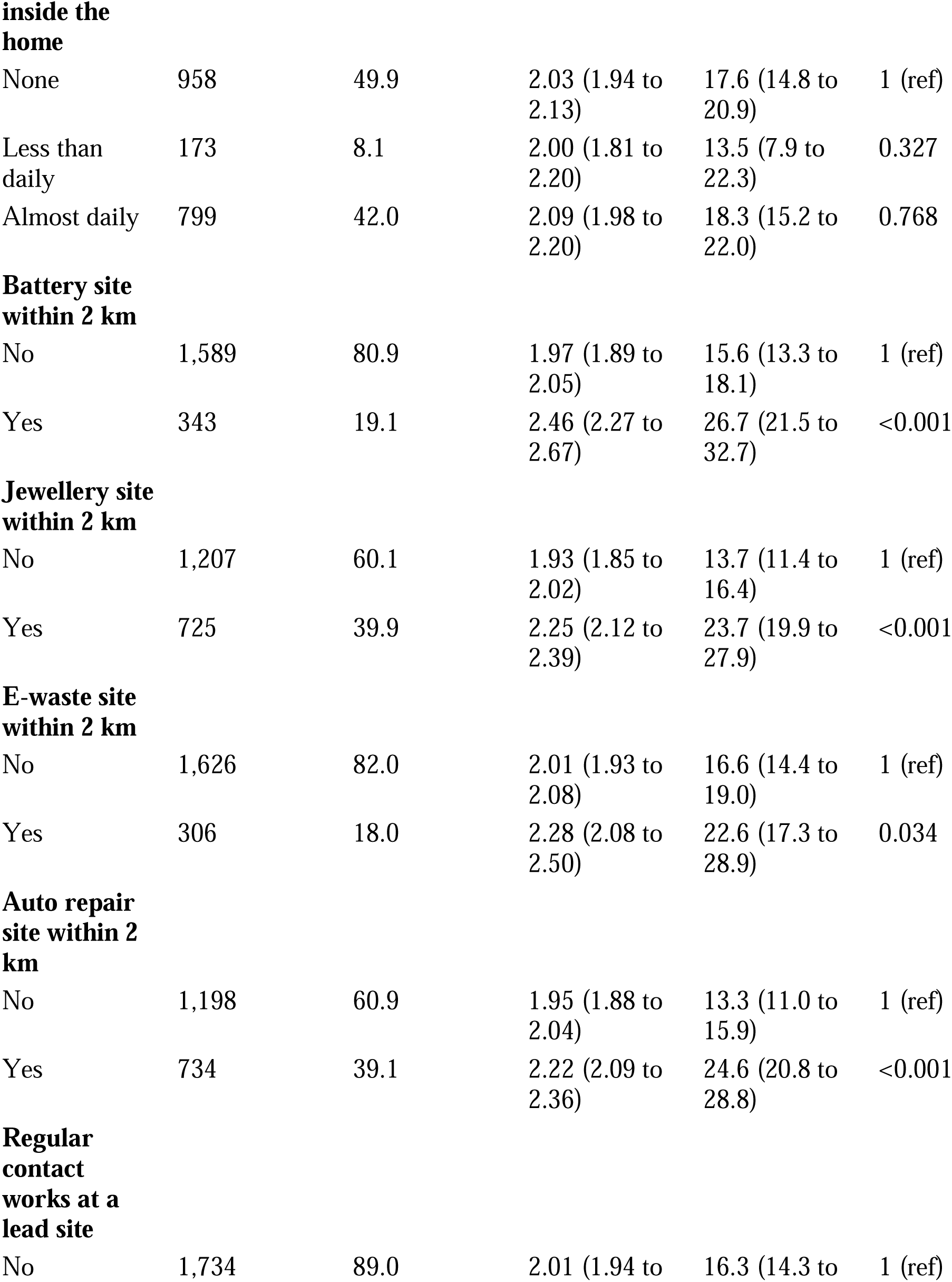
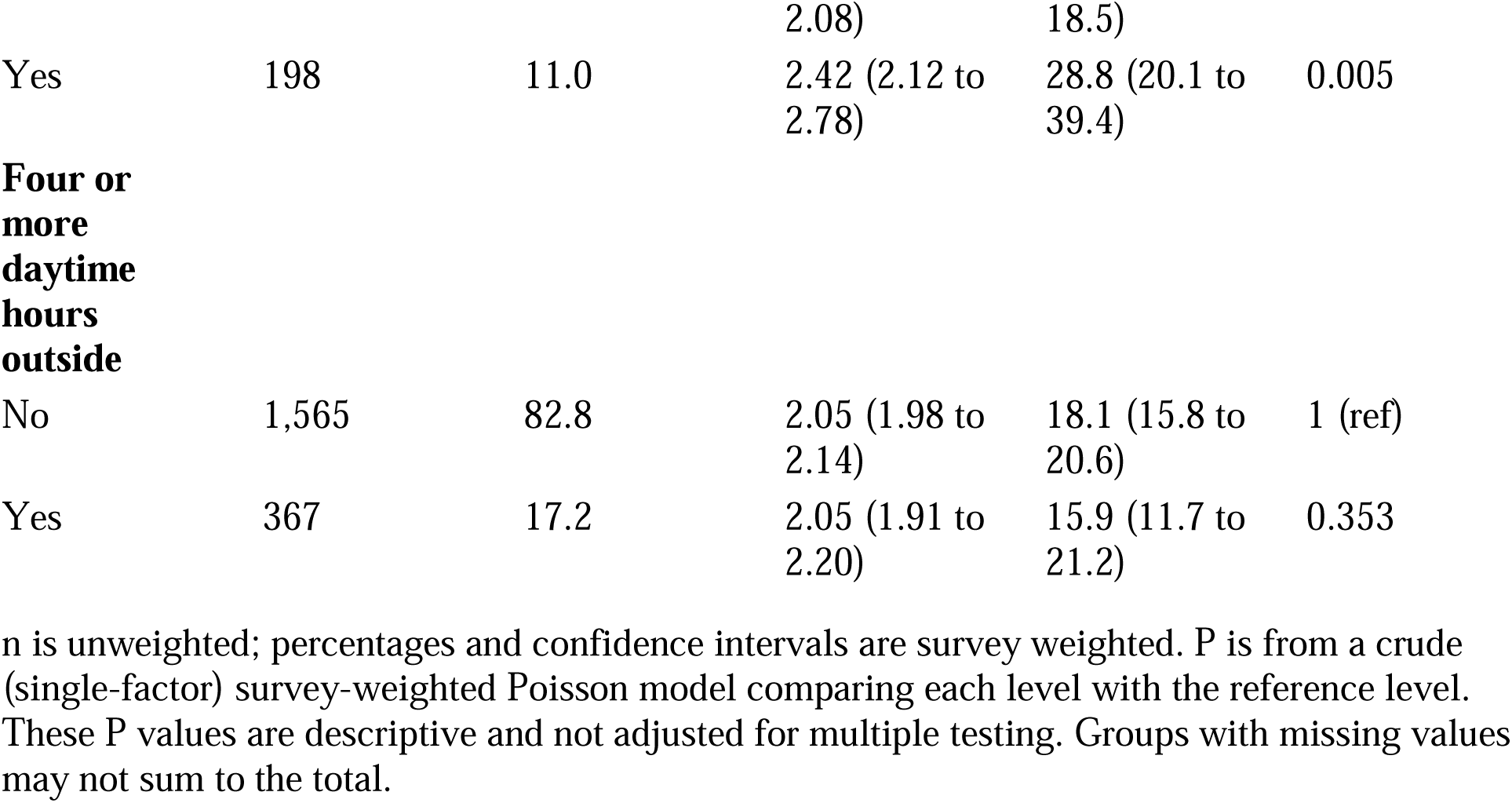
Characteristics of pregnant women with a valid blood test and blood lead by characteristic, Bangladesh MICS 2025 (n = 1,932).

### Levels and geography of blood lead

Among tested women, the geometric mean blood lead was 2.05 µg/dL (95% CI 1.99 to 2.12), and the 95th percentile was 5.83 µg/dL (Table 2, Fig 1). In all, 17.7% (15.7 to 19.9) had 3.5 µg/dL or more, 7.6% (6.1 to 9.3) had 5 µg/dL or more and 1.1% had 10 µg/dL or more. With response weights, these were 17.7% (15.8 to 19.8) and 7.6% (6.3 to 9.2) (S11 Table). Response weights ranged from 0.06 to 5.2 times their mean. Only six women had a lead value at the lowest reported level, and excluding them barely changed the geometric mean (2.07 µg/dL; S15 Table).

**Table 2.**
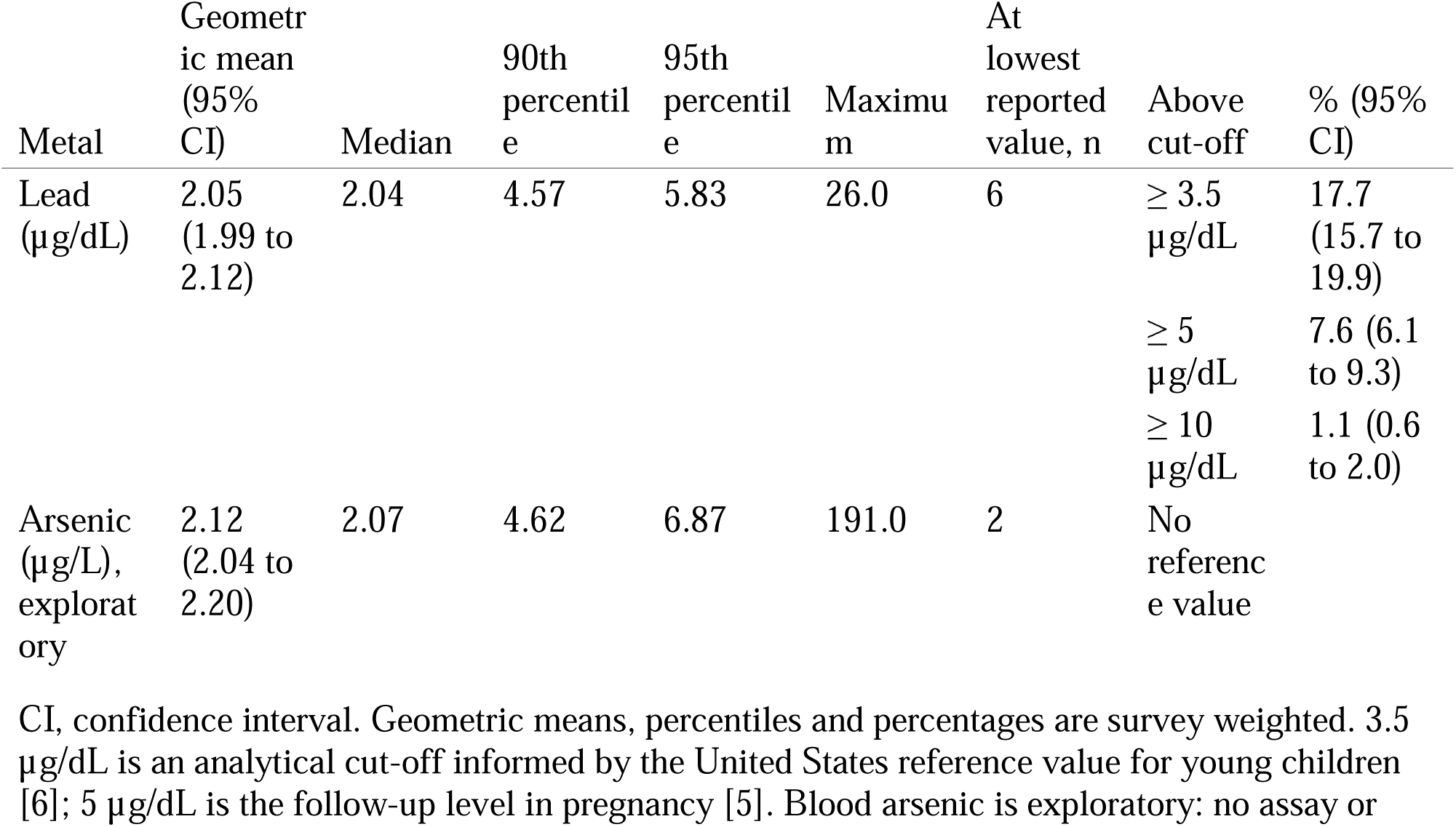
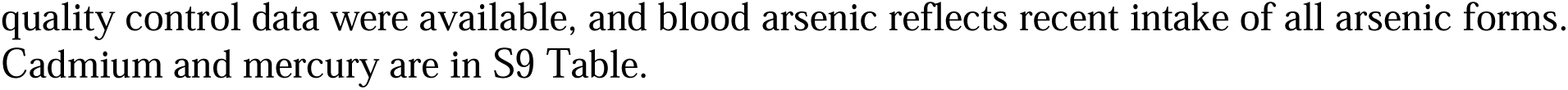
Blood lead and blood arsenic in pregnant women, Bangladesh MICS 2025 (n = 1,932).

| Metal | Geometric mean (95% CI) | Median | 90th percentile | 95th percentile | Maximum | At lowest reported value, n | Above cut-off | % (95% CI) |
| --- | --- | --- | --- | --- | --- | --- | --- | --- |
| Lead ($\mu\text{g/dL}$) | 2.05 (1.99 to 2.12) | 2.04 | 4.57 | 5.83 | 26.0 | 6 | $\geq 3.5$ $\mu\text{g/dL}$ | 17.7 (15.7 to 19.9) |
| | | | | | | | $\geq 5$ $\mu\text{g/dL}$ | 7.6 (6.1 to 9.3) |
| | | | | | | | $\geq 10$ $\mu\text{g/dL}$ | 1.1 (0.6 to 2.0) |
| Arsenic ($\mu\text{g/L}$), exploratory | 2.12 (2.04 to 2.20) | 2.07 | 4.62 | 6.87 | 191.0 | 2 | No reference value | |

CI, confidence interval. Geometric means, percentiles and percentages are survey weighted. 3.5 $\mu\text{g/dL}$ is an analytical cut-off informed by the United States reference value for young children [6]; 5 $\mu\text{g/dL}$ is the follow-up level in pregnancy [5]. Blood arsenic is exploratory: no assay or
quality control data were available, and blood arsenic reflects recent intake of all arsenic forms. Cadmium and mercury are in S9 Table.

**Fig 1.**
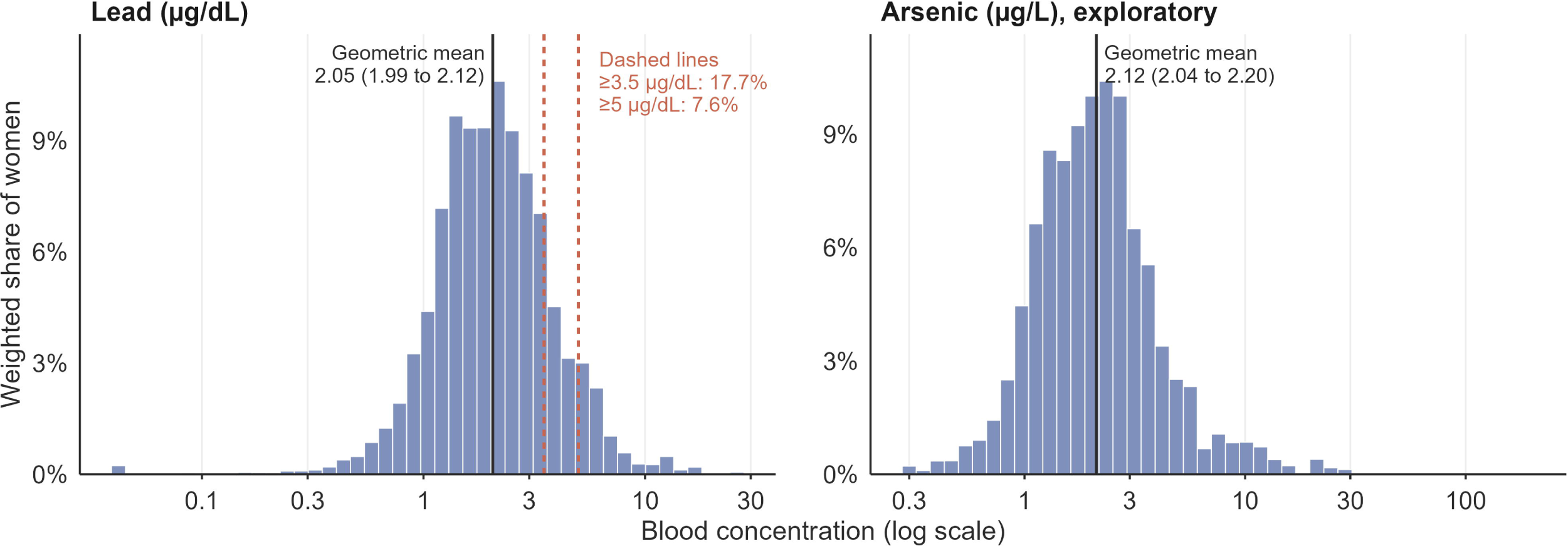
Distribution of blood lead and blood arsenic in pregnant women, Bangladesh MICS 2025. Survey-weighted histograms on a log scale (n = 1,932). Solid lines mark the geometric mean with its 95% confidence interval. Dashed lines mark 3.5 µg/dL, an analytical cut-off informed by the United States reference value for young children, and 5 µg/dL, the follow-up level in pregnancy. The weighted share of women at or above each cut-off is printed beside the lines. Arsenic is exploratory. Bars at the far left show results at the lowest reported value.

Raised lead was more common in urban (27.0%, 22.6 to 31.7) than rural women (13.8%, 11.8 to 16.2). It rose from 10.0% in the poorest to 30.7% in the richest wealth quintile (Table 1). Dhaka division stood apart, with 41.2% (35.5 to 47.0) at 3.5 µg/dL or more. Elsewhere the division figures ranged from 3.8% in Khulna to 15.6% in Sylhet.

The district map placed the highest levels in and around Dhaka (Fig 2A). District intervals were wide, reflecting 10 to 60 women per district (S8 Table). Greater Dhaka contributed 112 tested women from 85 clusters; 75 had 3.5 µg/dL or more and 47 had 5 µg/dL or more (S12 Table). There, 65.2% (55.4 to 73.9) of women had 3.5 µg/dL or more and 39.8% (30.0 to 50.6) had 5 µg/dL or more. The geometric mean was 4.30 µg/dL, against 1.79 µg/dL outside Dhaka division. Greater Dhaka held 12.1% (10.5 to 13.9) of pregnant women. It held 44.6% (38.1 to 51.2) of those at 3.5 µg/dL or more and 63.6% (53.5 to 72.7) of those at 5 µg/dL or more (Fig 2B). With response weights, these shares were 48.1% and 67.0% (S11 Table).

**Fig 2.**
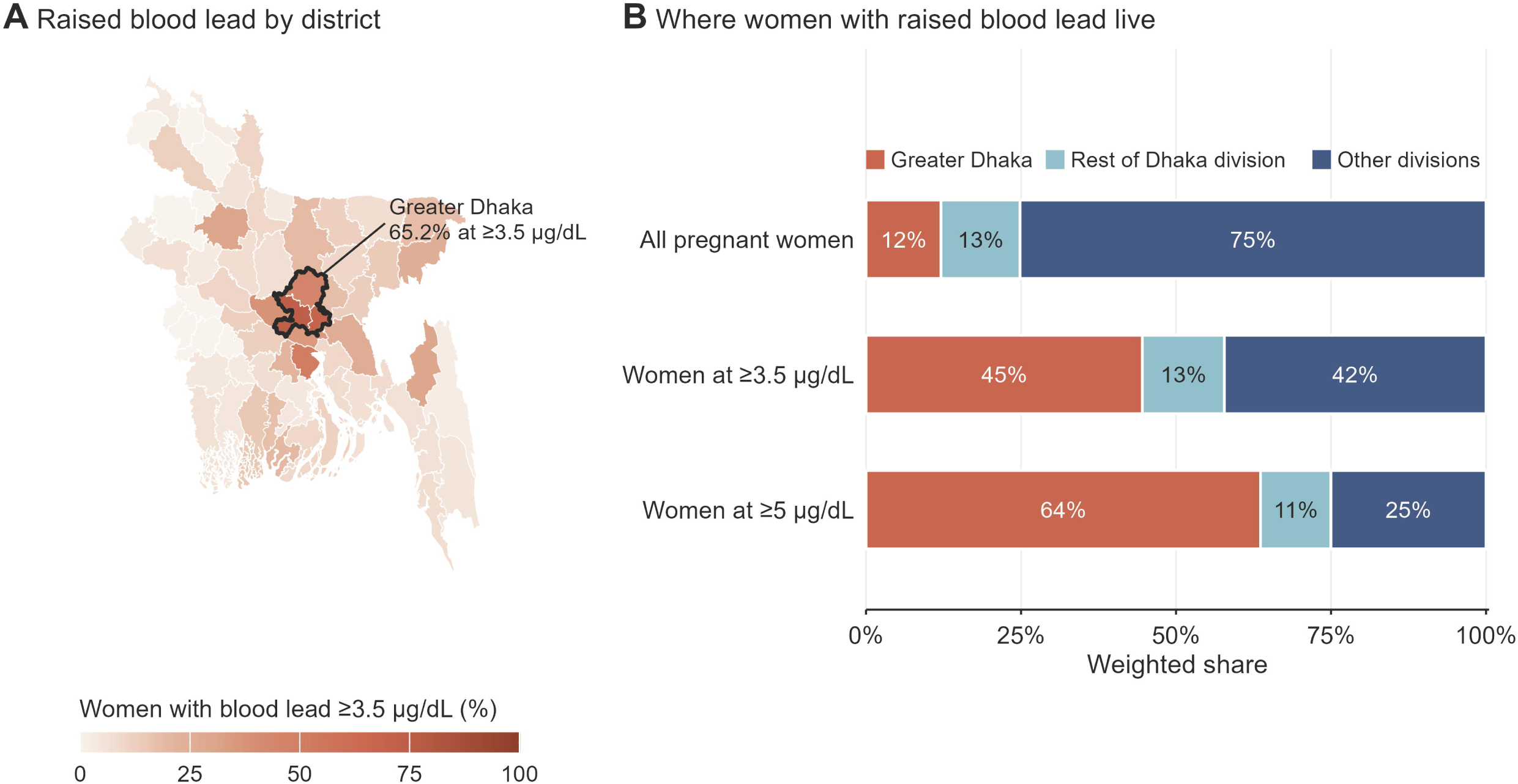
Where pregnant women with raised blood lead live. (A) Survey-weighted percentage of pregnant women with blood lead of 3.5 µg/dL or more by district, with city corporations merged into their districts. The colour scale runs from 0 to 100%, with colour steps concentrated over the observed range of 0 to 73%. The black outline marks greater Dhaka (Dhaka, Gazipur and Narayanganj), a grouping defined after inspecting the district estimates. District samples range from 10 to 60 women; S8 Table gives each district’s 95% confidence interval. (B) Weighted share of all pregnant women, and of women with raised lead, living in greater Dhaka, the rest of Dhaka division and the other divisions. S12 Table gives confidence intervals for these shares.

Arsenic, cadmium and mercury results are exploratory (Table 2, S2 Fig, S9 Table). The geometric mean of blood arsenic was 2.12 µg/L. Blood arsenic rose with source water arsenic. Against water below 10 ppb, the geometric mean ratio was 1.47 (1.13 to 1.91) at 10 to 49 ppb. It was 3.17 (2.44 to 4.11, P<0.001) at 50 ppb or more (S5 Table). Cadmium reached 1 µg/L in 24.5% of women, and mercury reached the 5.8 µg/L benchmark in 2.2% (S9 Table).

### Factors associated with raised blood lead

In crude analyses, raised lead was linked with wealth, urban residence, Dhaka division and every reported nearby site (Table 3). After adjustment in Model 1, Dhaka division kept a prevalence ratio of 2.80 (1.73 to 4.54, P<0.001) against Barishal. The richest quintile kept a ratio of 1.76 (1.15 to 2.71, P=0.010). Higher secondary education was linked with lower prevalence (0.66, 0.46 to 0.94, P=0.020). Of the exposure items, only a nearby automobile repair site remained associated (1.52, 1.07 to 2.16, P=0.021). No clear association was observed for sindoor (1.00, P=0.993), surma (1.06, P=0.653) or almost daily indoor smoking (1.09, P=0.515). The same held for a nearby battery site (1.13, 0.85 to 1.51, P=0.401) and contact with a lead site worker (1.18, 0.86 to 1.61, P=0.311).

**Table 3.**
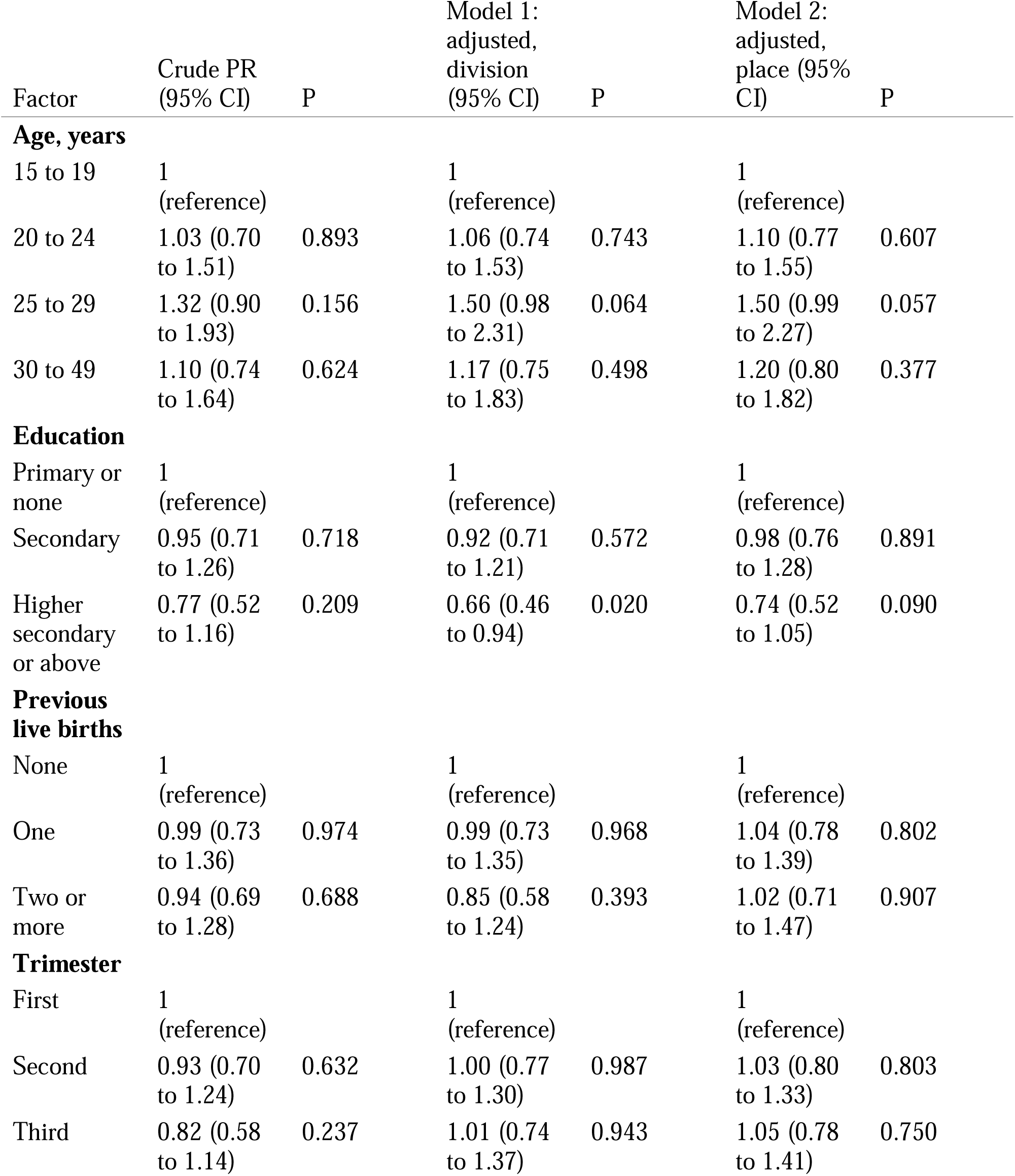
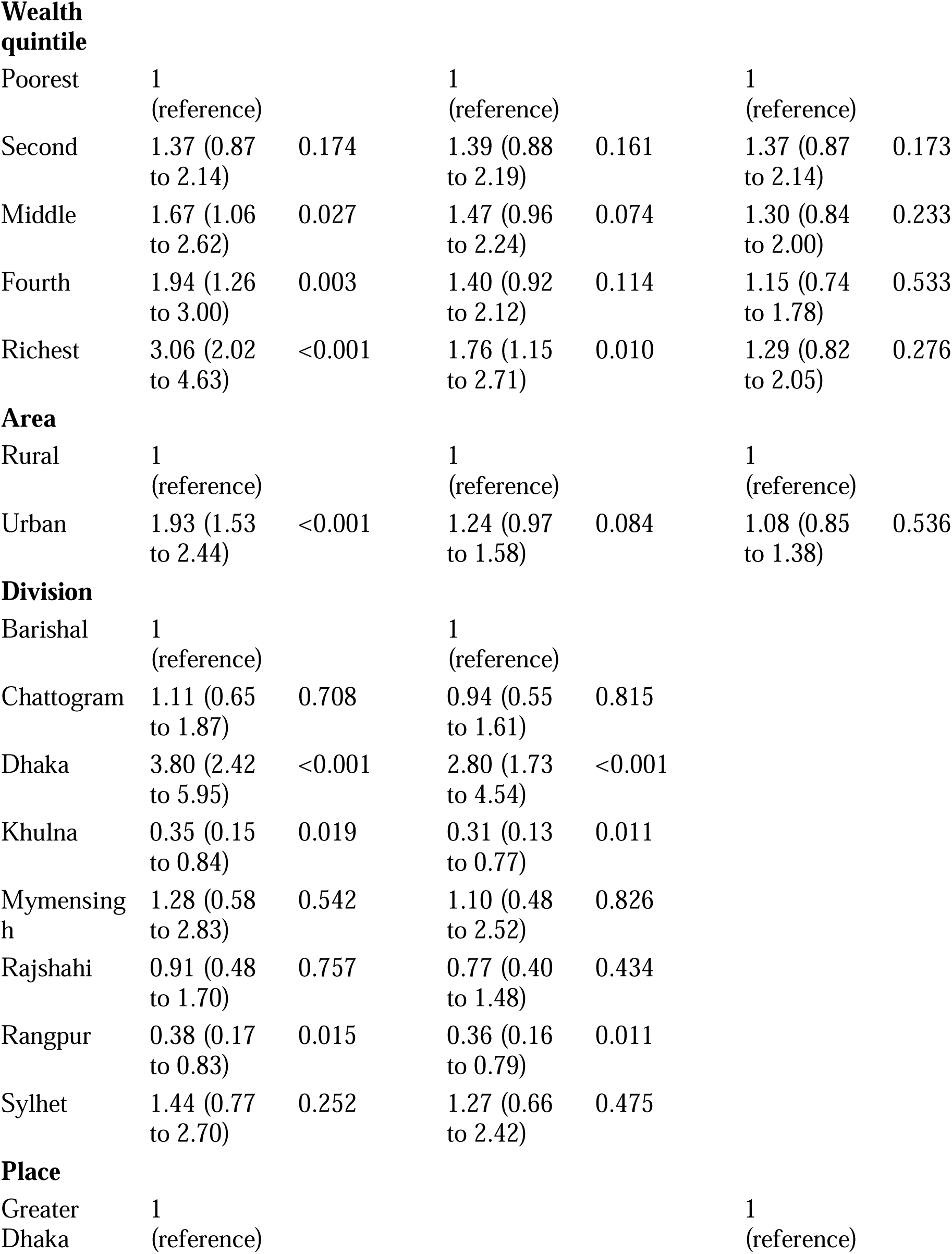
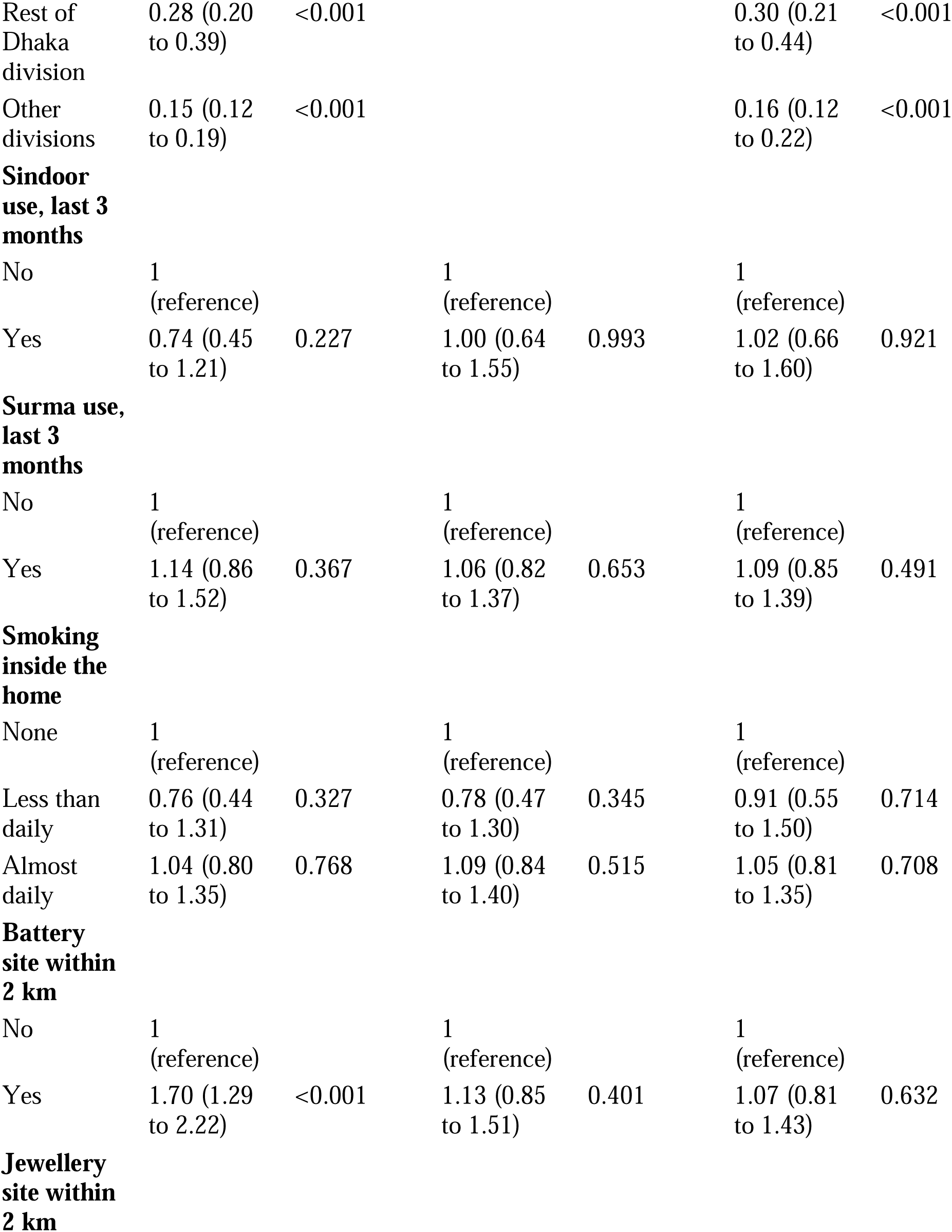
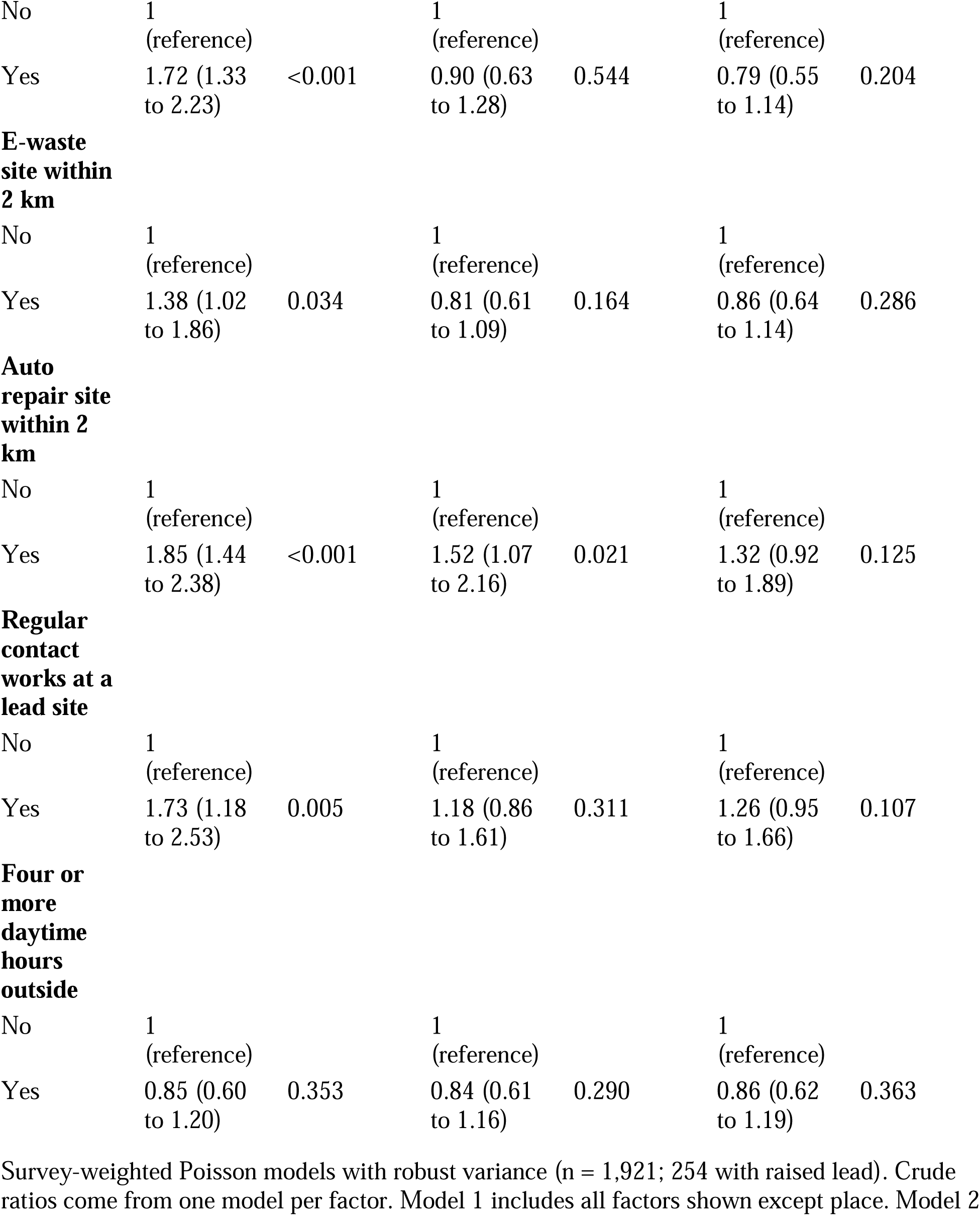
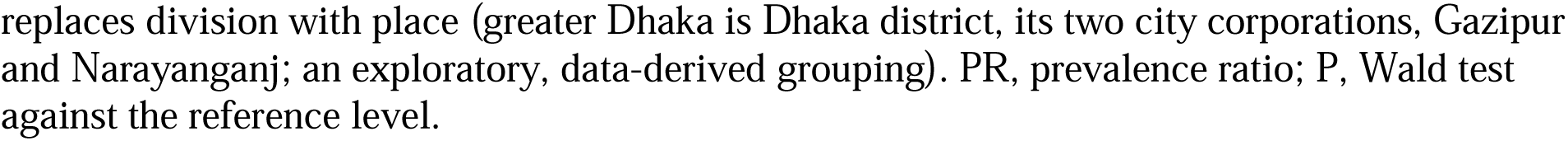
Prevalence ratios for blood lead of 3.5 µg/dL or more in pregnant women, Bangladesh MICS 2025.

| Factor | Crude PR<br>(95% CI) | P | Model 1:<br>adjusted,<br>division<br>(95% CI) | P | Model 2:<br>adjusted,<br>place (95%<br>CI) | P |
| --- | --- | --- | --- | --- | --- | --- |
| **Age, years** |  |  |  |  |  |  |
| 15 to 19 | 1<br>(reference) |  | 1<br>(reference) |  | 1<br>(reference) |  |
| 20 to 24 | 1.03 (0.70<br>to 1.51) | 0.893 | 1.06 (0.74<br>to 1.53) | 0.743 | 1.10 (0.77<br>to 1.55) | 0.607 |
| 25 to 29 | 1.32 (0.90<br>to 1.93) | 0.156 | 1.50 (0.98<br>to 2.31) | 0.064 | 1.50 (0.99<br>to 2.27) | 0.057 |
| 30 to 49 | 1.10 (0.74<br>to 1.64) | 0.624 | 1.17 (0.75<br>to 1.83) | 0.498 | 1.20 (0.80<br>to 1.82) | 0.377 |
| **Education** |  |  |  |  |  |  |
| Primary or<br>none | 1<br>(reference) |  | 1<br>(reference) |  | 1<br>(reference) |  |
| Secondary | 0.95 (0.71<br>to 1.26) | 0.718 | 0.92 (0.71<br>to 1.21) | 0.572 | 0.98 (0.76<br>to 1.28) | 0.891 |
| Higher<br>secondary<br>or above | 0.77 (0.52<br>to 1.16) | 0.209 | 0.66 (0.46<br>to 0.94) | 0.020 | 0.74 (0.52<br>to 1.05) | 0.090 |
| **Previous**<br>**live births** |  |  |  |  |  |  |
| None | 1<br>(reference) |  | 1<br>(reference) |  | 1<br>(reference) |  |
| One | 0.99 (0.73<br>to 1.36) | 0.974 | 0.99 (0.73<br>to 1.35) | 0.968 | 1.04 (0.78<br>to 1.39) | 0.802 |
| Two or<br>more | 0.94 (0.69<br>to 1.28) | 0.688 | 0.85 (0.58<br>to 1.24) | 0.393 | 1.02 (0.71<br>to 1.47) | 0.907 |
| **Trimester** |  |  |  |  |  |  |
| First | 1<br>(reference) |  | 1<br>(reference) |  | 1<br>(reference) |  |
| Second | 0.93 (0.70<br>to 1.24) | 0.632 | 1.00 (0.77<br>to 1.30) | 0.987 | 1.03 (0.80<br>to 1.33) | 0.803 |
| Third | 0.82 (0.58<br>to 1.14) | 0.237 | 1.01 (0.74<br>to 1.37) | 0.943 | 1.05 (0.78<br>to 1.41) | 0.750 |
| **Wealth quintile** |  |  |  |  |  |  |
| Poorest | 1<br>(reference) |  | 1<br>(reference) |  | 1<br>(reference) |  |
| Second | 1.37 (0.87<br>to 2.14) | 0.174 | 1.39 (0.88<br>to 2.19) | 0.161 | 1.37 (0.87<br>to 2.14) | 0.173 |
| Middle | 1.67 (1.06<br>to 2.62) | 0.027 | 1.47 (0.96<br>to 2.24) | 0.074 | 1.30 (0.84<br>to 2.00) | 0.233 |
| Fourth | 1.94 (1.26<br>to 3.00) | 0.003 | 1.40 (0.92<br>to 2.12) | 0.114 | 1.15 (0.74<br>to 1.78) | 0.533 |
| Richest | 3.06 (2.02<br>to 4.63) | <0.001 | 1.76 (1.15<br>to 2.71) | 0.010 | 1.29 (0.82<br>to 2.05) | 0.276 |
| **Area** |  |  |  |  |  |  |
| Rural | 1<br>(reference) |  | 1<br>(reference) |  | 1<br>(reference) |  |
| Urban | 1.93 (1.53<br>to 2.44) | <0.001 | 1.24 (0.97<br>to 1.58) | 0.084 | 1.08 (0.85<br>to 1.38) | 0.536 |
| **Division** |  |  |  |  |  |  |
| Barishal | 1<br>(reference) |  | 1<br>(reference) |  |  |  |
| Chattogram | 1.11 (0.65<br>to 1.87) | 0.708 | 0.94 (0.55<br>to 1.61) | 0.815 |  |  |
| Dhaka | 3.80 (2.42<br>to 5.95) | <0.001 | 2.80 (1.73<br>to 4.54) | <0.001 |  |  |
| Khulna | 0.35 (0.15<br>to 0.84) | 0.019 | 0.31 (0.13<br>to 0.77) | 0.011 |  |  |
| Mymensingh | 1.28 (0.58<br>to 2.83) | 0.542 | 1.10 (0.48<br>to 2.52) | 0.826 |  |  |
| Rajshahi | 0.91 (0.48<br>to 1.70) | 0.757 | 0.77 (0.40<br>to 1.48) | 0.434 |  |  |
| Rangpur | 0.38 (0.17<br>to 0.83) | 0.015 | 0.36 (0.16<br>to 0.79) | 0.011 |  |  |
| Sylhet | 1.44 (0.77<br>to 2.70) | 0.252 | 1.27 (0.66<br>to 2.42) | 0.475 |  |  |
| **Place** |  |  |  |  |  |  |
| Greater Dhaka | 1<br>(reference) |  |  |  | 1<br>(reference) |  |
| Rest of<br>Dhaka<br>division | 0.28 (0.20<br>to 0.39) | <0.001 |  |  | 0.30 (0.21<br>to 0.44) | <0.001 |
| Other<br>divisions | 0.15 (0.12<br>to 0.19) | <0.001 |  |  | 0.16 (0.12<br>to 0.22) | <0.001 |
| **Sindoor**<br>**use, last 3**<br>**months** |  |  |  |  |  |  |
| No | 1<br>(reference) |  | 1<br>(reference) |  | 1<br>(reference) |  |
| Yes | 0.74 (0.45<br>to 1.21) | 0.227 | 1.00 (0.64<br>to 1.55) | 0.993 | 1.02 (0.66<br>to 1.60) | 0.921 |
| **Surma use,**<br>**last 3**<br>**months** |  |  |  |  |  |  |
| No | 1<br>(reference) |  | 1<br>(reference) |  | 1<br>(reference) |  |
| Yes | 1.14 (0.86<br>to 1.52) | 0.367 | 1.06 (0.82<br>to 1.37) | 0.653 | 1.09 (0.85<br>to 1.39) | 0.491 |
| **Smoking**<br>**inside the**<br>**home** |  |  |  |  |  |  |
| None | 1<br>(reference) |  | 1<br>(reference) |  | 1<br>(reference) |  |
| Less than<br>daily | 0.76 (0.44<br>to 1.31) | 0.327 | 0.78 (0.47<br>to 1.30) | 0.345 | 0.91 (0.55<br>to 1.50) | 0.714 |
| Almost<br>daily | 1.04 (0.80<br>to 1.35) | 0.768 | 1.09 (0.84<br>to 1.40) | 0.515 | 1.05 (0.81<br>to 1.35) | 0.708 |
| **Battery**<br>**site within**<br>**2 km** |  |  |  |  |  |  |
| No | 1<br>(reference) |  | 1<br>(reference) |  | 1<br>(reference) |  |
| Yes | 1.70 (1.29<br>to 2.22) | <0.001 | 1.13 (0.85<br>to 1.51) | 0.401 | 1.07 (0.81<br>to 1.43) | 0.632 |
| **Jewellery**<br>**site within**<br>**2 km** |  |  |  |  |  |  |
| No | 1<br>(reference) |  | 1<br>(reference) |  | 1<br>(reference) |  |
| Yes | 1.72 (1.33<br>to 2.23) | <0.001 | 0.90 (0.63<br>to 1.28) | 0.544 | 0.79 (0.55<br>to 1.14) | 0.204 |
| **E-waste**<br>**site within**<br>**2 km** |  |  |  |  |  |  |
| No | 1<br>(reference) |  | 1<br>(reference) |  | 1<br>(reference) |  |
| Yes | 1.38 (1.02<br>to 1.86) | 0.034 | 0.81 (0.61<br>to 1.09) | 0.164 | 0.86 (0.64<br>to 1.14) | 0.286 |
| **Auto**<br>**repair site**<br>**within 2**<br>**km** |  |  |  |  |  |  |
| No | 1<br>(reference) |  | 1<br>(reference) |  | 1<br>(reference) |  |
| Yes | 1.85 (1.44<br>to 2.38) | <0.001 | 1.52 (1.07<br>to 2.16) | 0.021 | 1.32 (0.92<br>to 1.89) | 0.125 |
| **Regular**<br>**contact**<br>**works at a**<br>**lead site** |  |  |  |  |  |  |
| No | 1<br>(reference) |  | 1<br>(reference) |  | 1<br>(reference) |  |
| Yes | 1.73 (1.18<br>to 2.53) | 0.005 | 1.18 (0.86<br>to 1.61) | 0.311 | 1.26 (0.95<br>to 1.66) | 0.107 |
| **Four or**<br>**more**<br>**daytime**<br>**hours**<br>**outside** |  |  |  |  |  |  |
| No | 1<br>(reference) |  | 1<br>(reference) |  | 1<br>(reference) |  |
| Yes | 0.85 (0.60<br>to 1.20) | 0.353 | 0.84 (0.61<br>to 1.16) | 0.290 | 0.86 (0.62<br>to 1.19) | 0.363 |

Survey-weighted Poisson models with robust variance (n = 1,921; 254 with raised lead). Crude ratios come from one model per factor. Model 1 includes all factors shown except place. Model 2
replaces division with place (greater Dhaka is Dhaka district, its two city corporations, Gazipur and Narayanganj; an exploratory, data-derived grouping). PR, prevalence ratio; P, Wald test against the reference level.

Adding place attenuated most of the remaining differences (Model 2). Compared with greater Dhaka, prevalence was lower in the rest of Dhaka division (0.30, 0.21 to 0.44, P<0.001) and in other divisions (0.16, 0.12 to 0.22, P<0.001). With place in the model, the richest quintile ratio fell to 1.29 (0.82 to 2.05, P=0.276). The automobile repair site ratio fell to 1.32 (0.92 to 1.89, P=0.125). The wealth association weakened step by step as place was added (Fig 3A). The richest quintile ratio was 3.06 (2.02 to 4.63, P<0.001) in the wealth-only model and 1.69 (1.12 to 2.53, P=0.012) after urban residence and division. It was 1.09 (0.72 to 1.67, P=0.678) after adding greater Dhaka and 0.93 (0.60 to 1.46, P=0.759) with district fixed effects. The intervals remain wide, so a modest wealth effect within places cannot be excluded.

**Fig 3.**
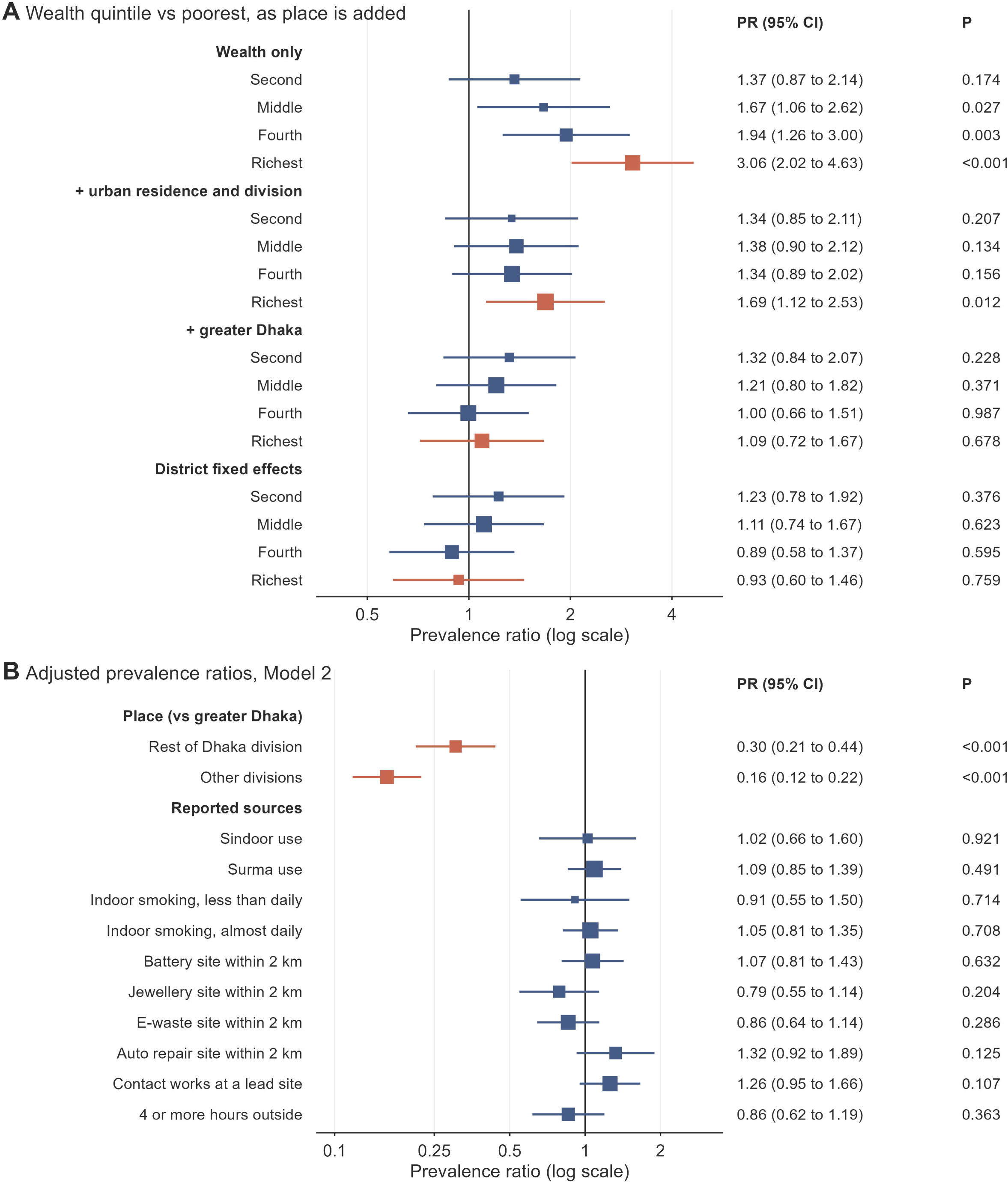
Prevalence ratios for blood lead of 3.5 µg/dL or more. (A) Wealth quintiles against the poorest in four survey-weighted Poisson models. The models are wealth only, then adding urban residence and division, then adding greater Dhaka, and finally district fixed effects in place of division. (B) Adjusted prevalence ratios from Model 2 (Table 3). Squares are sized by the inverse variance of each estimate, and lines show 95% confidence intervals on a log scale. P values are from Wald tests. Coloured squares mark the richest quintile in A and place in B.

Outside greater Dhaka, no wealth, urban or exposure factor showed a clear gradient (180 women with raised lead). Response weights left Model 1 almost unchanged; for example, the Dhaka division ratio was 2.81 (1.74 to 4.54) (S4 Table). In the extended model, households using polluting cooking fuel (0.46, P<0.001) or tube wells (0.56, P<0.001) had lower prevalence. Both mark life outside greater Dhaka, where 84% used polluting fuel and 80% tube wells, against 18% and 15% inside it. Household soil lead was weakly associated with raised blood lead (1.74 per log unit, 0.94 to 3.21, P=0.077). For the 5 µg/dL outcome, prevalence outside Dhaka division was 0.08 (0.04 to 0.14, P<0.001) times that in greater Dhaka. Higher secondary education was linked with lower prevalence (0.40, 0.21 to 0.77, P=0.006). Linear models of log lead gave the same pattern (S3 Table).

The site items overlapped moderately (correlations 0.40 to 0.64), but no variance inflation factor exceeded 3.1 (S14 Table). A simpler model gave the same place contrasts (0.28, 0.20 to 0.40, and 0.16, 0.12 to 0.21) and a richest quintile ratio of 1.31 (0.84 to 2.05).

### Links with child lead and haemoglobin

In 540 households, both the pregnant woman and a child aged 12 to 59 months had valid lead results (S7 Table). The woman was the child’s mother in 454 of them. Women with a tested child had slightly lower lead than other women (geometric mean 1.98 against 2.08 µg/dL), and fewer lived in greater Dhaka (7.3% against 13.8%). After adjustment for place and wealth, raised lead did not differ between them (PR 0.96, 0.73 to 1.25, P=0.742; S13 Table).

The child’s geometric mean was 4.57 µg/dL against 1.98 µg/dL for the woman, a ratio of 2.30 (2.15 to 2.47). The child had more lead than the woman in 486 of 540 pairs. Lead in women and children correlated moderately (Spearman 0.37, P<0.001; Fig 4). Each log unit of woman’s lead was associated with 0.24 (0.13 to 0.35, P<0.001) log units of child lead after adjustment. Adding household soil lead barely changed this (0.23, P<0.001). Where the woman had 3.5 µg/dL or more, 67.5% (53.7 to 78.8) of children had 5 µg/dL or more. Where she had less, 39.3% (33.9 to 44.9) did, an adjusted prevalence ratio of 1.50 (1.16 to 1.96, P=0.002). Results were similar for mother and child pairs (1.51, 1.15 to 1.98), with the child’s weight (1.58, 1.22 to 2.04) and with response weights (1.52, 1.16 to 1.99).

**Fig 4.**
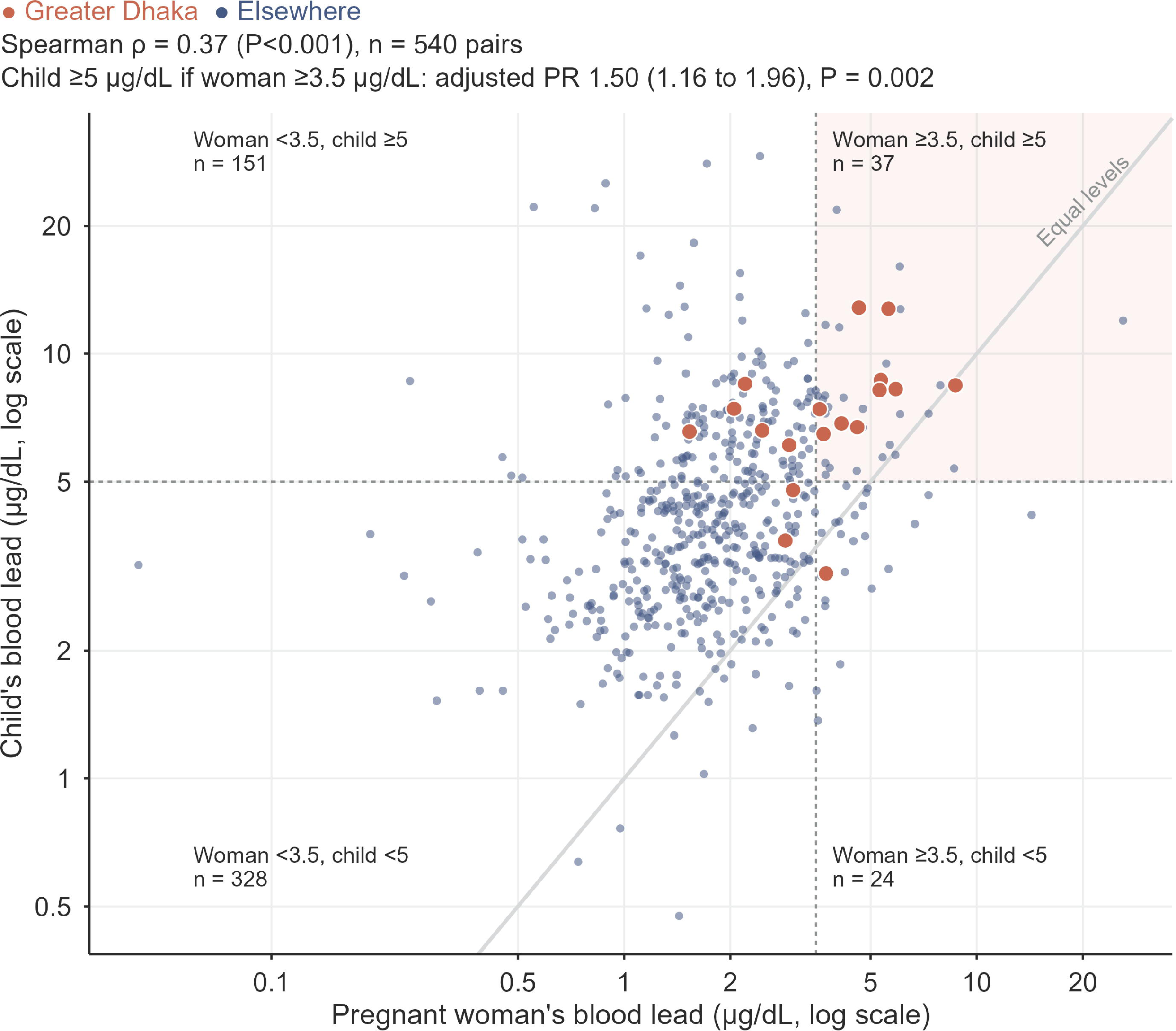
Blood lead in pregnant women and in a child aged 12 to 59 months of the same household. Each point is one household (n = 540); larger coloured points are in greater Dhaka. The grey diagonal marks equal levels. Dashed lines mark 3.5 µg/dL for the woman and 5 µg/dL for the child, and the shaded area marks households where both are raised. Counts in each quarter are unweighted. The Spearman correlation and the adjusted prevalence ratio are from survey-weighted Stata models (S7 Table).

As a marker, however, a woman’s lead of 3.5 µg/dL or more performed poorly (Table 4). It identified 22.7% (16.4 to 30.4) of children at 5 µg/dL or more; 151 of these 188 children lived with a woman below the cut-off. Specificity was 91.6% (87.3 to 94.6). The positive predictive value was 67.5% (53.7 to 78.8) and the negative predictive value 60.7% (55.1 to 66.1).

**Table 4.** A woman’s blood lead of 3.5 µg/dL or more as a marker of raised blood lead in a child aged 12 to 59 months of the same household (540 pairs).

| Measure | Child $\geq 5$ µg/dL:<br>n/N | % (95% CI) | Child $\geq 3.5$<br>µg/dL: n/N | % (95% CI) |
| --- | --- | --- | --- | --- |
| Sensitivity:<br>woman $\geq 3.5$<br>among children<br>above the cut-off | 37/188 | 22.7 (16.4 to<br>30.4) | 51/324 | 18.8 (14.2 to<br>24.4) |
| Specificity:<br>woman $< 3.5$<br>among children<br>below the cut-off | 328/352 | 91.6 (87.3 to<br>94.6) | 206/216 | 94.1 (88.2 to<br>97.1) |
| Positive<br>predictive value:<br>child above the<br>cut-off if woman<br>$\geq 3.5$ | 37/61 | 67.5 (53.7 to<br>78.8) | 51/61 | 86.6 (74.4 to<br>93.5) |
| Negative<br>predictive value:<br>child below the<br>cut-off if woman<br>$< 3.5$ | 328/479 | 60.7 (55.1 to<br>66.1) | 206/479 | 36.2 (31.3 to<br>41.5) |

n/N are unweighted counts; percentages and 95% confidence intervals are survey weighted with the woman's blood weight. Results apply to households with a pregnant woman and a tested child.

Anaemia affected 53.2% of pregnant women. It did not differ between women with and without raised lead (51.0% against 53.6%; adjusted PR 0.93, 0.80 to 1.09, P=0.396). Haemoglobin was not associated with log lead (difference -0.01 g/dL, -0.12 to 0.10, P=0.849). In the exploratory model with all four metals, each log unit of blood arsenic was associated with lower haemoglobin (difference -0.18 g/dL, -0.29 to -0.06, P=0.002; S6 Table).

## Discussion

### Main findings

To our knowledge, this is the first analysis of national data on blood lead in pregnant women in Bangladesh beyond the survey report. About one in six tested pregnant women had blood lead at or above the 3.5 µg/dL analytical cut-off, and one in 13 had 5 µg/dL or more. Raised lead varied sharply by place. Greater Dhaka held one in eight pregnant women but almost two thirds of those at 5 µg/dL or more. The crude wealth association was markedly attenuated after adjustment for place. No clear association was observed with sindoor, surma or indoor smoking. Lead in a pregnant woman was associated with lead in the young child of her household, but most exposed children lived with women below the cut-off. Lead was not clearly associated with anaemia.

### Levels of exposure

Our national figure of 7.6% at 5 µg/dL or more is much lower than the 31% found in an earlier study in rural central Bangladesh [8]. Rural women in our sample had only 4.9% at that level. Part of the difference may reflect the national campaign against lead chromate in turmeric after 2019 [10]. MICS did not measure turmeric or other foods, so this remains a hypothesis. The two studies also differ in the districts covered and possibly in laboratory methods. Our median of 2.0 µg/dL is below the 3.4 µg/dL of pregnant women in the United Kingdom in the early 1990s [22]. A national median hides very different conditions, however, as the greater Dhaka results show. Because no level of lead is known to be safe, the full distribution matters as much as any cut-off [2].

Children aged 12 to 59 months had more than twice the lead of the pregnant women in their homes. This fits the greater uptake and hand to mouth behaviour of young children. Blood lead in pregnancy may also be lowered by the rise in plasma volume. Lead released from the mother’s bone still reaches the fetus [4]. Women’s blood levels may therefore understate fetal exposure from long-term stores.

### Lead and place

The concentration in greater Dhaka is the main new finding. Dhaka, Gazipur and Narayanganj form the country’s main industrial region. In 2000, Dhaka schoolchildren had a mean blood lead of 15 µg/dL, when the city’s air lead was among the highest in the world [12]. A decade later, children in urban Dhaka still had higher levels than rural children, and living near industry predicted high levels [13]. Studies in urban Bangladesh have also found raised air lead from industry [23]. Our data cannot name the source in greater Dhaka. Reported nearby battery sites did not explain the excess, and the item relies on what women know of their neighbourhood. The weak link with automobile repair sites fell further within districts. It is more likely a marker of urban industrial neighbourhoods than a source in itself.

The greater Dhaka grouping was defined after seeing the data. It rests on 112 women in 85 clusters, so its estimates are uncertain. Its share of women with raised lead was stable with response weights, but it needs confirmation in new data.

Wealth is usually linked with lower lead exposure. In our data, the crude gradient ran the other way, with three times the prevalence in the richest quintile. Most of that association was attenuated once place was held fixed. Among tested women, 34% of the richest quintile lived in greater Dhaka, against under 1% of the poorest. Exposure there may be ambient and shared rather than tied to household poverty. Place may also stand for other unmeasured factors, so attenuation does not show what drives the gradient. A study of pregnant women in the United Kingdom also found higher lead with more education [22]. In our data, education was protective once place was held fixed. Targeting lead programmes by poverty alone may miss many exposed pregnant women in Bangladesh.

Sindoor and surma can contain lead. South Asian women in the United Kingdom who bought such cosmetics from ethnic shops had higher blood lead [24]. In our national sample, no clear association was observed for either practice. Products may differ in lead content, and our measure recorded use but not brand. Indoor smoking was also not clearly related to lead, although it was reported in about half of homes.

### Shared household exposure

Lead in pregnant women tracked lead in young children of the same household. Children of women with raised lead were 1.5 times as likely to have 5 µg/dL or more. The link did not depend on household soil lead, which suggests shared sources in food, dust, air or water. It was similar for mothers and their own children and with different weights. As a screening marker, however, maternal lead missed more than three in four children at 5 µg/dL or more. Maternal testing may flag some high-risk households, but it cannot replace testing of children or exposure assessment. The pair results apply to households with a pregnant woman and a young child. Women in these households had similar adjusted lead to other women.

### Lead, haemoglobin and the other metals

Lead was not clearly associated with haemoglobin or anaemia, and the estimates were close to null. MICS lacks data on iron status, infection and haemoglobinopathies. These are major causes of anaemia in Bangladesh, where thalassaemia and high groundwater iron matter more than iron deficiency alone [25]. We therefore cannot exclude a small effect of lead. Even among Dhaka children with much higher lead, the correlation with haemoglobin was weak [13]. The report’s table of anaemia by lead status should not be read as evidence of a causal link [17].

The survey report did not present arsenic, cadmium or mercury because of concerns about detection limits and precision [17]. We had no assay or quality control data for these metals, so we treat them as exploratory. Blood arsenic tripled at the highest source water levels, consistent with blood arsenic as a marker of recent intake from water [26]. This supports the ranking of arsenic values, not their absolute accuracy. The arsenic link with lower haemoglobin needs confirmation. The mercury benchmark refers to methylmercury, while the measured values are probably total mercury. The 2.2% above it therefore does not show excess methylmercury exposure.

### Implications

Our design cannot test screening programmes. The findings support confirmatory surveillance and source investigation in greater Dhaka, and studies of the feasibility and yield of targeted antenatal lead testing. Clinical guidance recommends follow-up, source investigation and nutritional advice at 5 µg/dL or more [5,27]. Because maternal lead identified few exposed children, child-focused testing and exposure assessment remain needed. Source investigation should look beyond the home to industrial emissions, dust and food supply chains. Soil remediation near an abandoned battery recycling site in Bangladesh modestly lowered children’s blood lead [28]. Similar action could follow where surveillance finds clusters of high results.

### Strengths and limitations

The study used venous blood in a national probability sample, and we reproduced the published report exactly. Linked child and soil results allowed household comparisons that clinic studies cannot make. Several limitations apply. Over a quarter of eligible women gave no valid sample, and refusal was more common among richer women and in Dhaka division. The blood weight corrects for differences in response between strata, and response weights within strata left the estimates almost unchanged. Bias remains possible if testing depended on lead level itself, and its direction is unknown. The design was cross-sectional, and the exposure items were self-reported and did not cover food, spices, cookware or occupation in detail. We defined greater Dhaka after seeing the district map, so its estimates are exploratory. District samples were small. Only 540 women had a tested child, and few of these pairs were in greater Dhaka. Blood lead was measured once, during pregnancy, when plasma volume changes. Gestational age was self-reported, so trimester may be misclassified. We examined many factors, and single associations near the significance threshold may arise by chance. No assay, detection limit or quality control data were available for any metal. The survey report treated the lead results as valid, but the other metals remain exploratory.

## Conclusions

Raised blood lead was common among pregnant women in Bangladesh and varied sharply by place, with the highest prevalence in greater Dhaka. The crude association between household wealth and raised lead was markedly attenuated after adjustment for place. Maternal lead was associated with lead in young children of the same household, although most children with raised lead lived with women below the cut-off. These findings support source investigation in greater Dhaka and evaluation of targeted maternal and child lead assessment, interpreted in light of incomplete testing and the cross-sectional design.

## Data Availability

The MICS dataset analysed in this study is publicly available from the UNICEF MICS website (https://mics.unicef.org) upon registration. GPS cluster coordinates were obtained under separate data request, due to location-sensitivity and displacement protocols.

## Supporting information

**S1 Fig.**
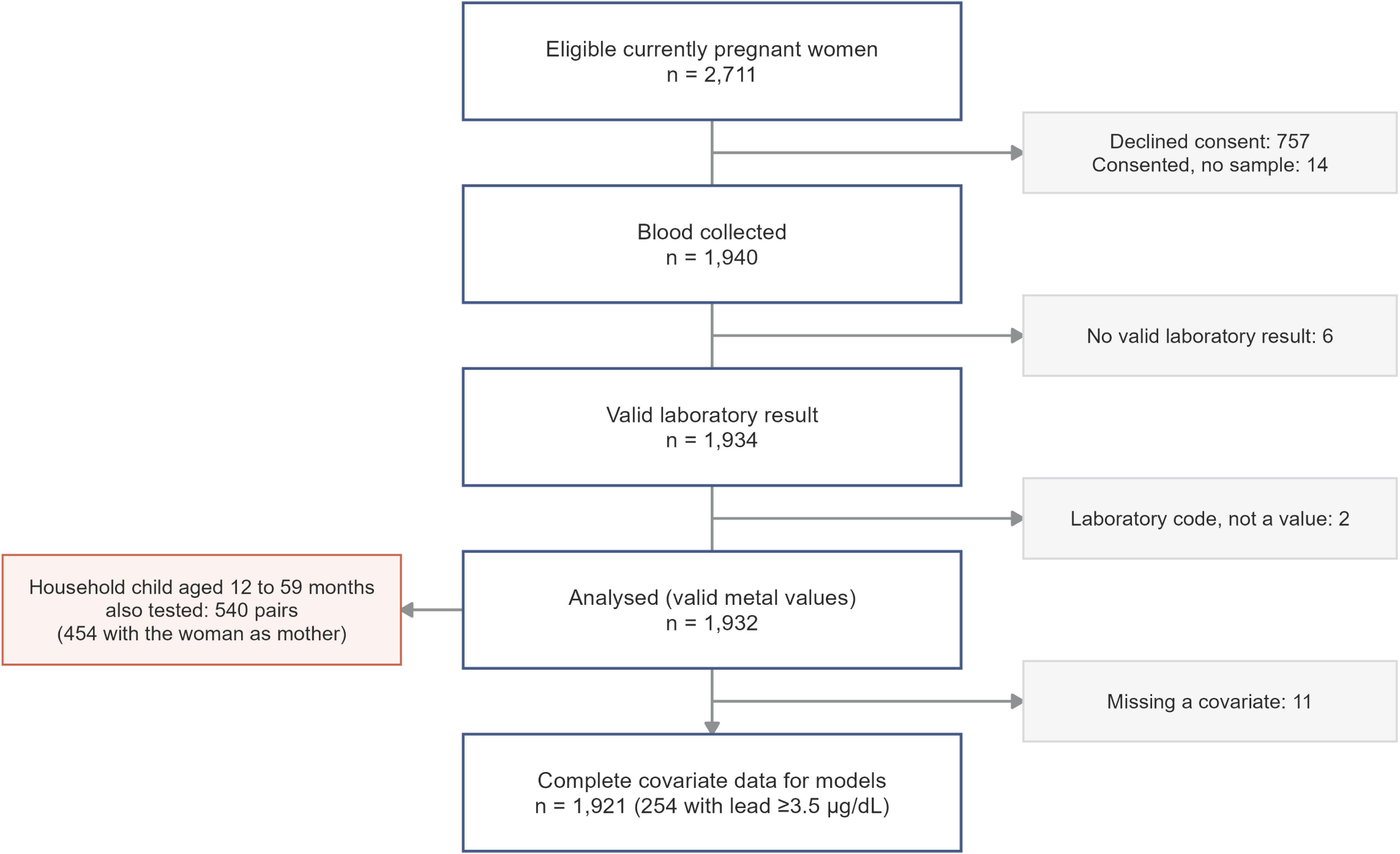
Participant flow, from eligible pregnant women to the analysis sample and the woman and child pairs.

**S2 Fig.**
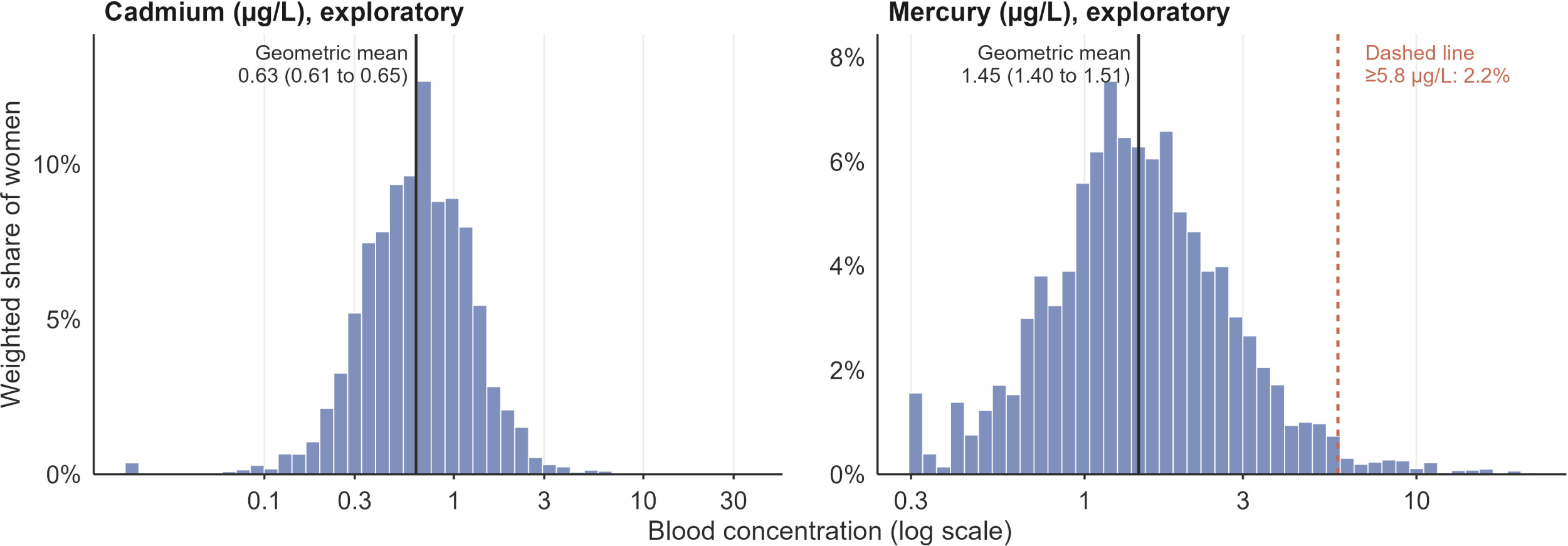
Distribution of blood cadmium and mercury in pregnant women (exploratory).

S1 File. Supplementary tables S1 to S15: response by characteristic; reproduction of the published report; geometric mean ratios; sensitivity analyses; other metals; haemoglobin; woman and child pairs; district estimates with confidence intervals; cadmium and mercury; response model and weight diagnostics; response-weighted estimates; greater Dhaka by component district; paired and unpaired women; model size and a simpler model; values at the lowest reported level.

S2 File. STROBE checklist.

## Declarations

### Author Contribution

Mohammad Khalid Hasan: Data curation, Methodology, Writing (original draft). Fahima Sharmin Hossain: Methodology, Writing (original draft). Md Khairul Islam: Methodology, Writing (review and editing). Ashika Sultana: Methodology, Writing (original draft). Md Ashifur Rahman: Methodology, Writing (original draft). Saad Mohammad Quaish: Data curation, Writing (original draft). Mohiuddin Sharif: Data curation, Writing (review and editing). Tanzin Naher: Data curation, Writing (review and editing). S. M. Ashraf Ul Hoque: Conceptualization, Formal analysis, Methodology, Project administration, Supervision, Validation, Visualization, Writing (review and editing). SMAUH is the guarantor and the corresponding author.

### Data availability

The Bangladesh MICS 2025 data sets are available free of charge from the UNICEF MICS website (https://mics.unicef.org/surveys). Users register, describe their intended use and download the data after approval. This study used the household, women’s and children under five files.

## References

1. Larsen B, Sánchez-Triana E. Global health burden and cost of lead exposure in children and adults: a health impact and economic modelling analysis. Lancet Planet Health. 2023;7(10):e831–e840. doi:10.1016/S2542-5196(23)00166-3

2. Lanphear BP, Hornung R, Khoury J, Yolton K, Baghurst P, Bellinger DC, et al. Low-level environmental lead exposure and children’s intellectual function: an international pooled analysis. Environ Health Perspect. 2005;113(7):894–9. doi:10.1289/ehp.7688

3. Hu H, Téllez-Rojo MM, Bellinger D, Smith D, Ettinger AS, Lamadrid-Figueroa H, et al. Fetal lead exposure at each stage of pregnancy as a predictor of infant mental development. Environ Health Perspect. 2006;114(11):1730–5. doi:10.1289/ehp.9067

4. Gulson BL, Mizon KJ, Palmer JM, Korsch MJ, Taylor AJ, Mahaffey KR. Blood lead changes during pregnancy and postpartum with calcium supplementation. Environ Health Perspect. 2004;112(15):1499–507. doi:10.1289/ehp.6548

5. Ettinger AS, Wengrovitz AG, editors. Guidelines for the identification and management of lead exposure in pregnant and lactating women. Atlanta: Centers for Disease Control and Prevention; 2010.

6. Ruckart PZ, Jones RL, Courtney JG, LeBlanc TT, Jackson W, Karwowski MP, et al. Update of the blood lead reference value: United States, 2021. MMWR Morb Mortal Wkly Rep. 2021;70(43):1509–12. doi:10.15585/mmwr.mm7043a4

7. Ericson B, Hu H, Nash E, Ferraro G, Sinitsky J, Taylor MP. Blood lead levels in low-income and middle-income countries: a systematic review. Lancet Planet Health. 2021;5(3):e145–e153. doi:10.1016/S2542-5196(20)30278-3

8. Forsyth JE, Saiful Islam M, Parvez SM, Raqib R, Sajjadur Rahman M, Marie Muehe E, et al. Prevalence of elevated blood lead levels among pregnant women and sources of lead exposure in rural Bangladesh: a case control study. Environ Res. 2018;166:1–9. doi:10.1016/j.envres.2018.04.019

9. Forsyth JE, Weaver KL, Maher K, Islam MS, Raqib R, Rahman M, et al. Sources of blood lead exposure in rural Bangladesh. Environ Sci Technol. 2019;53(19):11429–36. doi:10.1021/acs.est.9b00744

10. Forsyth JE, Baker M, Nurunnahar S, Islam S, Islam MS, Islam T, et al. Food safety policy enforcement and associated actions reduce lead chromate adulteration in turmeric across Bangladesh. Environ Res. 2023;232:116328. doi:10.1016/j.envres.2023.116328

11. Chowdhury KIA, Nurunnahar S, Kabir ML, Islam MT, Baker M, Islam MS, et al. Child lead exposure near abandoned lead acid battery recycling sites in a residential community in Bangladesh: risk factors and the impact of soil remediation on blood lead levels. Environ Res. 2021;194:110689. doi:10.1016/j.envres.2020.110689

12. Kaiser R, Henderson AK, Daley WR, Naughton M, Khan MH, Rahman M, et al. Blood lead levels of primary school children in Dhaka, Bangladesh. Environ Health Perspect. 2001;109(6):563–6. doi:10.1289/ehp.01109563

13. Mitra AK, Ahua E, Saha PK. Prevalence of and risk factors for lead poisoning in young children in Bangladesh. J Health Popul Nutr. 2012;30(4):404–9. doi:10.3329/jhpn.v30i4.13292

14. Gleason K, Shine JP, Shobnam N, Rokoff LB, Suchanda HS, Ibne Hasan MO, et al. Contaminated turmeric is a potential source of lead exposure for children in rural Bangladesh. J Environ Public Health. 2014;2014:730636. doi:10.1155/2014/730636

15. Lee MS, Eum KD, Golam M, Quamruzzaman Q, Kile ML, Mazumdar M, et al. Umbilical cord blood metal mixtures and birth size in Bangladeshi children. Environ Health Perspect. 2021;129(5):57006. doi:10.1289/EHP7502

16. Kippler M, Wagatsuma Y, Rahman A, Nermell B, Persson LÅ, Raqib R, et al. Environmental exposure to arsenic and cadmium during pregnancy and fetal size: a longitudinal study in rural Bangladesh. Reprod Toxicol. 2012;34(4):504–11. doi:10.1016/j.reprotox.2012.08.002

17. Bangladesh Bureau of Statistics; UNICEF Bangladesh. Progotir Pathey, Bangladesh Multiple Indicator Cluster Survey 2025, Survey Findings Report. Dhaka: BBS and UNICEF Bangladesh; 2026.

18. Khan S, Hancioglu A. Multiple Indicator Cluster Surveys: delivering robust data on children and women across the globe. Stud Fam Plann. 2019;50(3):279–86. doi:10.1111/sifp.12103

19. National Research Council. Toxicological effects of methylmercury. Washington, DC: National Academies Press; 2000.

20. Zou G. A modified Poisson regression approach to prospective studies with binary data. Am J Epidemiol. 2004;159(7):702–6. doi:10.1093/aje/kwh090

21. World Health Organization. Guideline on haemoglobin cutoffs to define anaemia in individuals and populations. Geneva: WHO; 2024.

22. Taylor CM, Golding J, Hibbeln J, Emond AM. Environmental factors predicting blood lead levels in pregnant women in the UK: the ALSPAC study. PLoS One. 2013;8(9):e72371. doi:10.1371/journal.pone.0072371

23. Woo MK, Young ES, Mostofa MG, Afroz S, Sharif Ibne Hasan MO, Quamruzzaman Q, et al. Lead in air in Bangladesh: exposure in a rural community with elevated blood lead concentrations among young children. Int J Environ Res Public Health. 2018;15(9):1947. doi:10.3390/ijerph15091947

24. Neelotpol S, Hay AW, Woolridge MW. Ethno-cultural risk of ante-natal lead exposure among South Asian women in the UK. Soc Sci Med. 2026;393:119034. doi:10.1016/j.socscimed.2026.119034

25. Merrill RD, Shamim AA, Ali H, Labrique AB, Schulze K, Christian P, et al. High prevalence of anemia with lack of iron deficiency among women in rural Bangladesh: a role for thalassemia and iron in groundwater. Asia Pac J Clin Nutr. 2012;21(3):416–24.

26. Hall M, Chen Y, Ahsan H, Slavkovich V, van Geen A, Parvez F, et al. Blood arsenic as a biomarker of arsenic exposure: results from a prospective study. Toxicology. 2006;225(2-3):225–33. doi:10.1016/j.tox.2006.06.010

27. World Health Organization. Guideline for the clinical management of exposure to lead. Geneva: WHO; 2021.

28. Rahman M, Sultana J, Sarker S, Hasan SS, Parvez SM, Das M, et al. Effectiveness of soil remediation intervention of abandoned used lead-acid battery recycling sites to reduce lead exposure among children: a three-arm pretest-posttest non-equivalent comparison group trial. Int J Hyg Environ Health. 2026;273:114756. doi:10.1016/j.ijheh.2026.114756

