## Supplementary material for "Blood lead and other metals among pregnant women in Bangladesh: national prevalence, geographic variation and household child exposure": S2_File_STROBE_checklist.docx

S2 File. STROBE checklist for cross-sectional studies

| Item | Recommendation | Where reported |
| --- | --- | --- |
| 1a | Study design in title or abstract | Abstract, Methods: “cross-sectional analysis” of a national survey |
| 1b | Informative, balanced abstract | Abstract |
| 2 | Scientific background and rationale | Introduction, paragraphs 1 to 3 |
| 3 | Objectives | Introduction, final paragraph |
| 4 | Key elements of study design | Methods, Study design and data |
| 5 | Setting, locations, dates of recruitment and data collection | Methods, Study design and data; Participants and blood testing (February to June 2025) |
| 6a | Eligibility criteria and selection of participants | Methods, Participants and blood testing; S1 Fig |
| 7 | Outcomes, exposures, predictors, confounders | Methods, Outcomes; Exposures and covariates |
| 8 | Data sources and measurement | Methods, Participants and blood testing; Outcomes; limits of laboratory documentation stated |
| 9 | Efforts to address bias | Methods, Missing data and sensitivity analyses (response weights, sensitivity models) |
| 10 | Study size | Methods, Study design and data (no sample size calculation; secondary analysis); S1 Fig |
| 11 | Handling of quantitative variables | Methods, Outcomes (log scale and cut-offs); Exposures and covariates |
| 12a | Statistical methods, including confounding control | Methods, Statistical analysis |
| 12b | Subgroups and interactions | Methods, Factors associated with raised blood lead (outside greater Dhaka; mother and child pairs) |
| 12c | Missing data | Methods, Missing data and sensitivity analyses; S1 Fig |
| 12d | Sampling strategy in analysis | Methods, Statistical analysis (weights, clusters, strata, subpopulation estimation) |
| 12e | Sensitivity analyses | Methods, Missing data and sensitivity analyses; S4, S11, S14, S15 Tables |
| 13a | Numbers at each stage | Results, Participants; S1 Fig |
| 13b | Reasons for non-participation | S1 Fig (declined consent, no sample, no valid result, laboratory code) |
| 13c | Flow diagram | S1 Fig |
| 14a | Characteristics of participants | Table 1; S1 Table |
| 14b | Missing data for each variable | Table 1 footnote; S1 Fig (11 women missing a covariate) |
| 15 | Outcome events or summary measures | Table 2; Results, Levels and geography of blood lead |
| 16a | Unadjusted and adjusted estimates with precision | Table 3; S3 and S4 Tables |
| 16b | Category boundaries | Methods, Outcomes; Table 1 |
| 16c | Absolute risk where relevant | Table 1 and Table 2 (weighted prevalence) |
| 17 | Other analyses | Results; Table 4; S5 to S15 Tables |
| 18 | Key results | Discussion, Main findings |
| 19 | Limitations | Discussion, Strengths and limitations |
| 20 | Interpretation | Discussion |
| 21 | Generalisability | Discussion, Strengths and limitations; Shared household exposure (target population of pair results) |
| 22 | Funding | Declarations |
