## Supplementary material for "Blood lead and other metals among pregnant women in Bangladesh: national prevalence, geographic variation and household child exposure": Supplement_Draft_v3.docx

S1 File. Supplementary tables

All estimates are survey weighted unless stated otherwise and were produced in Stata 17. Arsenic, cadmium and mercury results are exploratory.

### S1 Table. Eligible pregnant women and those with a valid blood test, by characteristic

| Characteristic | Eligible, n | Valid test, n | Valid test, % |
| --- | --- | --- | --- |
| **Age, years** |  |  |  |
| 15 to 19 | 686 | 487 | 71.0 |
| 20 to 24 | 860 | 596 | 69.3 |
| 25 to 29 | 632 | 448 | 70.9 |
| 30 to 49 | 533 | 403 | 75.6 |
| **Education** |  |  |  |
| Primary or none | 520 | 415 | 79.8 |
| Secondary | 1494 | 1093 | 73.2 |
| Higher secondary or above | 696 | 425 | 61.1 |
| **Previous live births** |  |  |  |
| None | 1071 | 712 | 66.5 |
| One | 898 | 658 | 73.3 |
| Two or more | 742 | 564 | 76.0 |
| **Trimester** |  |  |  |
| First | 806 | 580 | 72.0 |
| Second | 1107 | 811 | 73.3 |
| Third | 797 | 542 | 68.0 |
| **Wealth quintile** |  |  |  |
| Poorest | 551 | 453 | 82.2 |
| Second | 584 | 445 | 76.2 |
| Middle | 563 | 397 | 70.5 |
| Fourth | 564 | 394 | 69.9 |
| Richest | 449 | 245 | 54.6 |
| **Area** |  |  |  |
| Urban | 756 | 519 | 68.7 |
| Rural | 1955 | 1415 | 72.4 |
| **Division** |  |  |  |
| Barishal | 238 | 207 | 87.0 |
| Chattogram | 650 | 429 | 66.0 |
| Dhaka | 587 | 359 | 61.2 |
| Khulna | 349 | 248 | 71.1 |
| Mymensingh | 135 | 94 | 69.6 |
| Rajshahi | 259 | 199 | 76.8 |
| Rangpur | 298 | 250 | 83.9 |
| Sylhet | 195 | 148 | 75.9 |
| **Religion of household head** |  |  |  |
| Not Hindu | 2503 | 1781 | 71.2 |
| Hindu | 208 | 153 | 73.6 |
| **All** | 2711 | 1934 | 71.3 |

Unweighted counts. Trimester and education are missing for a few women, so groups may not sum to the total.

### S2 Table. Reproduction of published lead and anaemia estimates (MICS 2025 report Tables ATW.14.1S and ATW.15.1S)

| Estimate | This analysis | Published report |
| --- | --- | --- |
| Completed blood collections, n | 1940 | 1,940 |
| Blood lead below 3.5 µg/dL, % | 81.9 | 81.9 |
| 3.5 to 4.99 µg/dL, % | 10.1 | 10.1 |
| 5.0 to 9.99 µg/dL, % | 6.4 | 6.4 |
| 10 µg/dL or more, % | 1.1 | 1.1 |
| Lab reject or barcode mismatch, % | 0.5 | 0.5 |
| Mean blood lead, µg/dL | 2.51 | 2.51 |
| Blood lead 5 µg/dL or more, urban, % | 14.0 | 14.0 |
| Blood lead 5 µg/dL or more, rural, % | 4.9 | 4.9 |
| Blood lead 5 µg/dL or more, poorest to richest quintile, % | 2.3, 4.2, 7.0, 10.4, 15.0 | 2.3, 4.2, 7.0, 10.4, 15.0 |
| Mean haemoglobin, g/dL | 10.68 | 10.68 |
| Any anaemia, % (WHO 2024 trimester-specific cut-offs) | 53.2 | 53.1 |
| Blood lead 5 µg/dL or more, Dhaka division, % | 22.8 | 22.8 |

Report percentages for lead use all 1,940 completed collections as the denominator, with lab rejects as a separate category. The anaemia figure in this analysis excludes the six collections without a valid result.

### S3 Table. Adjusted geometric mean ratios for blood lead

| Factor | Model 1, division (95% CI) | P | Model 2, place (95% CI) | P |
| --- | --- | --- | --- | --- |
| Age 20 to 24 | 1.04 (0.95 to 1.15) | 0.369 | 1.05 (0.96 to 1.15) | 0.262 |
| Age 25 to 29 | 1.11 (0.99 to 1.25) | 0.085 | 1.11 (0.98 to 1.24) | 0.089 |
| Age 30 to 49 | 1.07 (0.94 to 1.21) | 0.297 | 1.07 (0.95 to 1.21) | 0.241 |
| Secondary education | 0.92 (0.85 to 1.00) | 0.040 | 0.92 (0.85 to 1.00) | 0.045 |
| Higher secondary or above | 0.80 (0.72 to 0.88) | <0.001 | 0.82 (0.74 to 0.90) | <0.001 |
| One previous birth | 0.98 (0.91 to 1.07) | 0.717 | 1.00 (0.92 to 1.08) | 1.000 |
| Two or more previous births | 0.95 (0.86 to 1.06) | 0.388 | 1.02 (0.92 to 1.13) | 0.726 |
| Second trimester | 0.96 (0.90 to 1.04) | 0.322 | 0.98 (0.92 to 1.05) | 0.597 |
| Third trimester | 1.03 (0.96 to 1.11) | 0.449 | 1.05 (0.98 to 1.13) | 0.172 |
| Wealth: second | 1.09 (1.00 to 1.19) | 0.046 | 1.09 (1.00 to 1.19) | 0.053 |
| Wealth: middle | 1.08 (0.99 to 1.19) | 0.085 | 1.06 (0.97 to 1.16) | 0.205 |
| Wealth: fourth | 1.11 (1.01 to 1.22) | 0.023 | 1.08 (0.99 to 1.19) | 0.086 |
| Wealth: richest | 1.22 (1.09 to 1.37) | <0.001 | 1.14 (1.02 to 1.28) | 0.022 |
| Urban | 1.06 (0.98 to 1.14) | 0.158 | 1.01 (0.94 to 1.09) | 0.764 |
| Chattogram | 0.94 (0.83 to 1.07) | 0.351 |  |  |
| Dhaka | 1.46 (1.29 to 1.66) | <0.001 |  |  |
| Khulna | 0.74 (0.64 to 0.85) | <0.001 |  |  |
| Mymensingh | 0.94 (0.78 to 1.13) | 0.494 |  |  |
| Rajshahi | 0.81 (0.69 to 0.95) | 0.008 |  |  |
| Rangpur | 0.77 (0.67 to 0.88) | <0.001 |  |  |
| Sylhet | 1.04 (0.89 to 1.22) | 0.609 |  |  |
| Rest of Dhaka division (vs greater Dhaka) |  |  | 0.56 (0.49 to 0.64) | <0.001 |
| Other divisions (vs greater Dhaka) |  |  | 0.43 (0.38 to 0.49) | <0.001 |
| Sindoor use | 0.98 (0.88 to 1.08) | 0.649 | 0.97 (0.87 to 1.08) | 0.584 |
| Surma use | 1.05 (0.98 to 1.13) | 0.166 | 1.08 (1.01 to 1.16) | 0.018 |
| Indoor smoking, less than daily | 0.96 (0.87 to 1.06) | 0.468 | 1.02 (0.92 to 1.12) | 0.704 |
| Indoor smoking, almost daily | 1.03 (0.97 to 1.10) | 0.307 | 1.03 (0.97 to 1.09) | 0.394 |
| Battery site within 2 km | 1.11 (1.00 to 1.22) | 0.041 | 1.10 (1.00 to 1.20) | 0.055 |
| Jewellery site within 2 km | 1.01 (0.92 to 1.10) | 0.817 | 0.97 (0.89 to 1.07) | 0.580 |
| E-waste site within 2 km | 0.95 (0.85 to 1.05) | 0.323 | 0.97 (0.88 to 1.07) | 0.516 |
| Auto repair site within 2 km | 1.03 (0.94 to 1.12) | 0.555 | 0.98 (0.90 to 1.06) | 0.565 |
| Regular contact works at a lead site | 1.05 (0.93 to 1.18) | 0.434 | 1.07 (0.96 to 1.19) | 0.202 |
| Four or more hours outside | 0.98 (0.91 to 1.05) | 0.542 | 0.98 (0.91 to 1.05) | 0.560 |

Survey-weighted linear models of log blood lead; ratios are exponentiated coefficients. Both models include all factors shown. n = 1,921. P is the Wald test for that term.

### S4 Table. Sensitivity analyses for blood lead of 3.5 µg/dL or more (adjusted prevalence ratios)

| Factor | Main model | P | Response weights | P | Extra household factors | P | District fixed effects | P | Soil subsample | P | Outcome 5 µg/dL or more, place model | P |
| --- | --- | --- | --- | --- | --- | --- | --- | --- | --- | --- | --- | --- |
| Age 20 to 24 | 1.06 (0.74 to 1.53) | 0.743 | 1.10 (0.75 to 1.60) | 0.631 | 0.97 (0.68 to 1.38) | 0.859 | 0.98 (0.71 to 1.35) | 0.894 | 1.83 (0.75 to 4.47) | 0.185 | 0.85 (0.50 to 1.45) | 0.550 |
| Age 25 to 29 | 1.50 (0.98 to 2.31) | 0.064 | 1.48 (0.95 to 2.31) | 0.080 | 1.37 (0.90 to 2.09) | 0.145 | 1.28 (0.90 to 1.84) | 0.170 | 2.38 (0.91 to 6.23) | 0.078 | 1.18 (0.62 to 2.23) | 0.621 |
| Age 30 to 49 | 1.17 (0.75 to 1.83) | 0.498 | 1.24 (0.79 to 1.94) | 0.358 | 0.97 (0.64 to 1.49) | 0.907 | 1.00 (0.66 to 1.50) | 0.991 | 1.74 (0.55 to 5.53) | 0.347 | 0.77 (0.40 to 1.50) | 0.447 |
| Secondary education | 0.92 (0.71 to 1.21) | 0.572 | 0.90 (0.69 to 1.18) | 0.436 | 1.07 (0.82 to 1.41) | 0.602 | 1.00 (0.77 to 1.31) | 0.977 | 0.47 (0.27 to 0.84) | 0.010 | 0.59 (0.38 to 0.93) | 0.023 |
| Higher secondary or above | 0.66 (0.46 to 0.94) | 0.020 | 0.65 (0.46 to 0.93) | 0.019 | 0.86 (0.60 to 1.24) | 0.423 | 0.77 (0.54 to 1.11) | 0.160 | 0.14 (0.05 to 0.40) | <0.001 | 0.40 (0.21 to 0.77) | 0.006 |
| One previous birth | 0.99 (0.73 to 1.35) | 0.968 | 0.96 (0.70 to 1.32) | 0.820 | 1.02 (0.76 to 1.36) | 0.912 | 1.13 (0.85 to 1.48) | 0.400 | 2.66 (0.71 to 10.01) | 0.148 | 1.22 (0.74 to 2.03) | 0.438 |
| Two or more previous births | 0.85 (0.58 to 1.24) | 0.393 | 0.82 (0.56 to 1.19) | 0.293 | 1.09 (0.75 to 1.57) | 0.660 | 1.14 (0.79 to 1.63) | 0.479 | 1.11 (0.26 to 4.77) | 0.889 | 0.82 (0.42 to 1.61) | 0.564 |
| Second trimester | 1.00 (0.77 to 1.30) | 0.987 | 0.98 (0.76 to 1.27) | 0.865 | 1.02 (0.80 to 1.31) | 0.848 | 1.00 (0.79 to 1.28) | 0.977 | 1.23 (0.66 to 2.28) | 0.511 | 0.77 (0.50 to 1.19) | 0.241 |
| Third trimester | 1.01 (0.74 to 1.37) | 0.943 | 1.01 (0.74 to 1.38) | 0.949 | 0.98 (0.72 to 1.32) | 0.886 | 0.98 (0.74 to 1.30) | 0.879 | 0.87 (0.44 to 1.73) | 0.693 | 1.00 (0.65 to 1.56) | 0.985 |
| Wealth: second | 1.39 (0.88 to 2.19) | 0.161 | 1.34 (0.84 to 2.15) | 0.221 | 1.32 (0.84 to 2.08) | 0.231 | 1.24 (0.80 to 1.93) | 0.329 | 1.70 (0.76 to 3.82) | 0.198 | 1.85 (0.81 to 4.19) | 0.142 |
| Wealth: middle | 1.47 (0.96 to 2.24) | 0.074 | 1.42 (0.93 to 2.19) | 0.106 | 1.06 (0.68 to 1.67) | 0.786 | 1.15 (0.75 to 1.74) | 0.522 | 0.90 (0.39 to 2.05) | 0.797 | 1.84 (0.86 to 3.95) | 0.116 |
| Wealth: fourth | 1.40 (0.92 to 2.12) | 0.114 | 1.38 (0.90 to 2.11) | 0.134 | 0.72 (0.46 to 1.15) | 0.168 | 0.95 (0.62 to 1.46) | 0.813 | 1.01 (0.46 to 2.24) | 0.979 | 1.59 (0.71 to 3.55) | 0.259 |
| Wealth: richest | 1.76 (1.15 to 2.71) | 0.010 | 1.73 (1.11 to 2.70) | 0.015 | 0.70 (0.41 to 1.18) | 0.175 | 1.02 (0.63 to 1.66) | 0.930 | 1.15 (0.44 to 2.99) | 0.780 | 1.73 (0.76 to 3.91) | 0.191 |
| Urban | 1.24 (0.97 to 1.58) | 0.084 | 1.28 (1.00 to 1.63) | 0.046 | 0.97 (0.75 to 1.24) | 0.790 | 1.32 (1.01 to 1.74) | 0.042 | 1.28 (0.77 to 2.14) | 0.342 | 1.12 (0.74 to 1.69) | 0.604 |
| Chattogram | 0.94 (0.55 to 1.61) | 0.815 | 0.85 (0.50 to 1.46) | 0.566 | 0.87 (0.51 to 1.49) | 0.603 |  |  | 1.48 (0.56 to 3.93) | 0.429 |  |  |
| Dhaka | 2.80 (1.73 to 4.54) | <0.001 | 2.81 (1.74 to 4.54) | <0.001 | 2.27 (1.39 to 3.69) | <0.001 |  |  | 2.83 (1.06 to 7.54) | 0.037 |  |  |
| Khulna | 0.31 (0.13 to 0.77) | 0.011 | 0.29 (0.12 to 0.68) | 0.004 | 0.32 (0.13 to 0.79) | 0.014 |  |  | 0.31 (0.06 to 1.59) | 0.159 |  |  |
| Mymensingh | 1.10 (0.48 to 2.52) | 0.826 | 1.12 (0.47 to 2.70) | 0.795 | 0.93 (0.43 to 2.02) | 0.858 |  |  | 1.24 (0.29 to 5.37) | 0.771 |  |  |
| Rajshahi | 0.77 (0.40 to 1.48) | 0.434 | 0.74 (0.39 to 1.42) | 0.370 | 0.70 (0.37 to 1.34) | 0.279 |  |  | 0.96 (0.26 to 3.52) | 0.954 |  |  |
| Rangpur | 0.36 (0.16 to 0.79) | 0.011 | 0.36 (0.16 to 0.81) | 0.014 | 0.38 (0.17 to 0.85) | 0.018 |  |  | 0.22 (0.03 to 1.71) | 0.147 |  |  |
| Sylhet | 1.27 (0.66 to 2.42) | 0.475 | 1.23 (0.65 to 2.34) | 0.524 | 1.23 (0.64 to 2.33) | 0.533 |  |  | 1.22 (0.36 to 4.16) | 0.749 |  |  |
| Rest of Dhaka division (vs greater Dhaka) |  |  |  |  |  |  |  |  |  |  | 0.22 (0.12 to 0.41) | <0.001 |
| Other divisions (vs greater Dhaka) |  |  |  |  |  |  |  |  |  |  | 0.08 (0.04 to 0.14) | <0.001 |
| Sindoor use | 1.00 (0.64 to 1.55) | 0.993 | 0.99 (0.63 to 1.55) | 0.956 | 0.84 (0.41 to 1.74) | 0.646 | 1.15 (0.77 to 1.73) | 0.486 | 0.51 (0.13 to 2.02) | 0.337 | 1.06 (0.42 to 2.69) | 0.904 |
| Surma use | 1.06 (0.82 to 1.37) | 0.653 | 1.02 (0.78 to 1.33) | 0.902 | 1.08 (0.85 to 1.37) | 0.542 | 1.12 (0.88 to 1.44) | 0.364 | 0.62 (0.33 to 1.18) | 0.149 | 1.33 (0.91 to 1.94) | 0.137 |
| Indoor smoking, less than daily | 0.78 (0.47 to 1.30) | 0.345 | 0.80 (0.48 to 1.33) | 0.384 | 0.79 (0.48 to 1.31) | 0.370 | 1.02 (0.63 to 1.66) | 0.923 | 1.47 (0.78 to 2.78) | 0.235 | 0.54 (0.21 to 1.41) | 0.208 |
| Indoor smoking, almost daily | 1.09 (0.84 to 1.40) | 0.515 | 1.04 (0.80 to 1.34) | 0.770 | 1.09 (0.85 to 1.39) | 0.517 | 1.08 (0.84 to 1.39) | 0.554 | 1.02 (0.59 to 1.74) | 0.956 | 0.88 (0.60 to 1.27) | 0.484 |
| Battery site within 2 km | 1.13 (0.85 to 1.51) | 0.401 | 1.15 (0.87 to 1.52) | 0.328 | 1.17 (0.88 to 1.56) | 0.278 | 0.93 (0.68 to 1.27) | 0.647 | 1.54 (0.79 to 3.01) | 0.200 | 1.03 (0.63 to 1.68) | 0.910 |
| Jewellery site within 2 km | 0.90 (0.63 to 1.28) | 0.544 | 0.84 (0.59 to 1.21) | 0.350 | 0.81 (0.56 to 1.16) | 0.251 | 0.88 (0.62 to 1.23) | 0.451 | 0.84 (0.39 to 1.79) | 0.653 | 0.94 (0.52 to 1.71) | 0.850 |
| E-waste site within 2 km | 0.81 (0.61 to 1.09) | 0.164 | 0.77 (0.57 to 1.04) | 0.092 | 0.83 (0.63 to 1.10) | 0.202 | 0.94 (0.68 to 1.30) | 0.710 | 0.60 (0.29 to 1.26) | 0.180 | 0.79 (0.47 to 1.32) | 0.364 |
| Auto repair site within 2 km | 1.52 (1.07 to 2.16) | 0.021 | 1.56 (1.09 to 2.23) | 0.016 | 1.53 (1.08 to 2.16) | 0.016 | 1.36 (0.96 to 1.94) | 0.083 | 1.55 (0.70 to 3.40) | 0.278 | 1.19 (0.70 to 2.03) | 0.525 |
| Regular contact works at a lead site | 1.18 (0.86 to 1.61) | 0.311 | 1.16 (0.83 to 1.62) | 0.385 | 1.09 (0.82 to 1.46) | 0.549 | 1.20 (0.89 to 1.62) | 0.222 | 1.66 (0.75 to 3.67) | 0.207 | 1.20 (0.73 to 1.98) | 0.474 |
| Four or more hours outside | 0.84 (0.61 to 1.16) | 0.290 | 0.79 (0.56 to 1.10) | 0.164 | 0.79 (0.57 to 1.09) | 0.150 | 0.85 (0.61 to 1.18) | 0.326 | 0.98 (0.55 to 1.75) | 0.945 | 0.80 (0.46 to 1.38) | 0.426 |
| Household soil lead, per log unit |  |  |  |  |  |  |  |  | 1.74 (0.94 to 3.21) | 0.077 |  |  |
| Hindu household |  |  |  |  | 1.42 (0.74 to 2.73) | 0.286 |  |  |  |  |  |  |
| Polluting cooking fuel |  |  |  |  | 0.46 (0.33 to 0.64) | <0.001 |  |  |  |  |  |  |
| Tube well (vs piped) |  |  |  |  | 0.56 (0.40 to 0.79) | <0.001 |  |  |  |  |  |  |
| Other water source (vs piped) |  |  |  |  | 1.05 (0.64 to 1.72) | 0.859 |  |  |  |  |  |  |
| Collected in March (vs February) |  |  |  |  | 2.01 (1.16 to 3.48) | 0.012 |  |  |  |  |  |  |
| Collected in April |  |  |  |  | 1.60 (0.90 to 2.84) | 0.106 |  |  |  |  |  |  |
| Collected in May |  |  |  |  | 1.41 (0.79 to 2.50) | 0.246 |  |  |  |  |  |  |
| Collected in June |  |  |  |  | 2.21 (1.23 to 3.97) | 0.008 |  |  |  |  |  |  |

Main model is Model 1 of Table 3. Response weights divide the women’s design weight by the modelled probability of a valid test (S10 Table). Extra household factors adds religion, cooking fuel, water source and month of collection. District fixed effects replaces division with 64 district terms (estimates not shown). Soil subsample adds household soil lead for the 534 women with a valid household soil result and complete data. The 5 µg/dL model uses place instead of division because Mymensingh had no woman at that level. P is the Wald test for that term.

### S5 Table. Adjusted geometric mean ratios for blood arsenic, cadmium and mercury (exploratory)

| Factor | Arsenic | P | Cadmium | P | Mercury | P | Arsenic, water test subsample | P |
| --- | --- | --- | --- | --- | --- | --- | --- | --- |
| Age 20 to 24 | 1.00 (0.89 to 1.13) | 0.951 | 1.08 (0.98 to 1.18) | 0.125 | 1.03 (0.94 to 1.14) | 0.517 | 1.25 (0.95 to 1.63) | 0.106 |
| Age 25 to 29 | 1.05 (0.92 to 1.20) | 0.484 | 1.13 (1.00 to 1.26) | 0.044 | 1.16 (1.03 to 1.30) | 0.016 | 1.01 (0.75 to 1.37) | 0.931 |
| Age 30 to 49 | 1.05 (0.91 to 1.21) | 0.514 | 1.12 (0.99 to 1.27) | 0.075 | 1.20 (1.06 to 1.37) | 0.005 | 1.09 (0.80 to 1.50) | 0.571 |
| Secondary education | 0.99 (0.91 to 1.09) | 0.894 | 0.93 (0.86 to 1.00) | 0.056 | 1.05 (0.97 to 1.13) | 0.273 | 1.02 (0.82 to 1.27) | 0.841 |
| Higher secondary or above | 0.95 (0.85 to 1.06) | 0.341 | 0.96 (0.86 to 1.07) | 0.477 | 1.05 (0.95 to 1.16) | 0.369 | 0.94 (0.74 to 1.20) | 0.633 |
| One previous birth | 0.97 (0.88 to 1.08) | 0.609 | 0.98 (0.90 to 1.08) | 0.727 | 0.92 (0.84 to 1.00) | 0.041 | 1.01 (0.79 to 1.29) | 0.951 |
| Two or more previous births | 1.00 (0.88 to 1.14) | 0.964 | 0.98 (0.87 to 1.10) | 0.712 | 0.94 (0.84 to 1.04) | 0.223 | 1.05 (0.77 to 1.43) | 0.748 |
| Second trimester | 1.03 (0.94 to 1.11) | 0.554 | 0.92 (0.85 to 0.99) | 0.031 | 0.83 (0.77 to 0.90) | <0.001 | 1.12 (0.93 to 1.33) | 0.227 |
| Third trimester | 1.04 (0.95 to 1.15) | 0.366 | 0.94 (0.87 to 1.02) | 0.140 | 0.87 (0.80 to 0.96) | 0.003 | 1.14 (0.92 to 1.40) | 0.233 |
| Wealth: second | 1.01 (0.91 to 1.12) | 0.853 | 0.97 (0.89 to 1.07) | 0.575 | 1.04 (0.95 to 1.14) | 0.447 | 0.94 (0.75 to 1.20) | 0.635 |
| Wealth: middle | 1.09 (0.97 to 1.23) | 0.144 | 0.89 (0.80 to 0.99) | 0.026 | 1.12 (1.01 to 1.24) | 0.026 | 0.82 (0.64 to 1.04) | 0.100 |
| Wealth: fourth | 0.96 (0.85 to 1.09) | 0.539 | 0.78 (0.70 to 0.87) | <0.001 | 1.05 (0.94 to 1.18) | 0.362 | 0.82 (0.66 to 1.01) | 0.066 |
| Wealth: richest | 0.96 (0.83 to 1.12) | 0.614 | 0.76 (0.66 to 0.88) | <0.001 | 1.10 (0.96 to 1.26) | 0.171 | 0.77 (0.59 to 1.00) | 0.047 |
| Urban | 0.91 (0.83 to 1.00) | 0.041 | 0.95 (0.87 to 1.03) | 0.216 | 0.95 (0.87 to 1.03) | 0.179 | 0.95 (0.79 to 1.14) | 0.565 |
| Chattogram | 1.60 (1.41 to 1.83) | <0.001 | 0.79 (0.70 to 0.88) | <0.001 | 1.08 (0.95 to 1.23) | 0.213 | 0.94 (0.72 to 1.22) | 0.633 |
| Dhaka | 1.35 (1.21 to 1.52) | <0.001 | 0.90 (0.79 to 1.02) | 0.087 | 0.80 (0.70 to 0.91) | 0.001 | 1.05 (0.79 to 1.40) | 0.741 |
| Khulna | 1.47 (1.30 to 1.66) | <0.001 | 0.58 (0.51 to 0.66) | <0.001 | 0.94 (0.82 to 1.08) | 0.378 | 0.97 (0.69 to 1.37) | 0.885 |
| Mymensingh | 1.06 (0.90 to 1.25) | 0.502 | 1.22 (1.05 to 1.41) | 0.008 | 0.85 (0.70 to 1.04) | 0.107 | 0.85 (0.55 to 1.29) | 0.442 |
| Rajshahi | 1.21 (1.06 to 1.38) | 0.005 | 0.64 (0.56 to 0.74) | <0.001 | 0.84 (0.74 to 0.97) | 0.015 | 0.85 (0.63 to 1.13) | 0.265 |
| Rangpur | 1.20 (1.06 to 1.35) | 0.003 | 1.16 (1.03 to 1.32) | 0.019 | 0.94 (0.81 to 1.08) | 0.377 | 1.02 (0.74 to 1.41) | 0.897 |
| Sylhet | 1.47 (1.26 to 1.71) | <0.001 | 1.23 (1.08 to 1.41) | 0.002 | 1.50 (1.31 to 1.72) | <0.001 | 0.89 (0.58 to 1.36) | 0.587 |
| Sindoor use | 0.96 (0.86 to 1.09) | 0.555 | 0.90 (0.81 to 1.00) | 0.051 | 0.99 (0.91 to 1.09) | 0.906 |  |  |
| Surma use | 1.01 (0.93 to 1.08) | 0.875 | 1.02 (0.95 to 1.10) | 0.507 | 1.07 (0.99 to 1.15) | 0.098 |  |  |
| Indoor smoking, less than daily | 1.14 (1.02 to 1.28) | 0.018 | 1.02 (0.93 to 1.13) | 0.667 | 1.13 (1.03 to 1.25) | 0.011 |  |  |
| Indoor smoking, almost daily | 1.04 (0.96 to 1.12) | 0.311 | 1.09 (1.02 to 1.17) | 0.007 | 1.01 (0.94 to 1.08) | 0.744 |  |  |
| Battery site within 2 km | 1.04 (0.93 to 1.17) | 0.491 | 1.11 (0.99 to 1.25) | 0.085 | 1.00 (0.91 to 1.11) | 0.960 |  |  |
| Jewellery site within 2 km | 1.08 (0.97 to 1.20) | 0.181 | 0.96 (0.87 to 1.05) | 0.349 | 1.01 (0.92 to 1.11) | 0.789 |  |  |
| E-waste site within 2 km | 1.02 (0.91 to 1.14) | 0.734 | 0.91 (0.81 to 1.02) | 0.101 | 1.08 (0.97 to 1.19) | 0.144 |  |  |
| Auto repair site within 2 km | 0.91 (0.82 to 1.00) | 0.060 | 1.04 (0.95 to 1.14) | 0.352 | 0.91 (0.83 to 1.00) | 0.061 |  |  |
| Regular contact works at a lead site | 0.99 (0.89 to 1.10) | 0.852 | 1.00 (0.90 to 1.12) | 0.994 | 1.00 (0.90 to 1.11) | 0.960 |  |  |
| Four or more hours outside | 1.00 (0.91 to 1.10) | 0.948 | 0.94 (0.87 to 1.02) | 0.146 | 0.95 (0.87 to 1.03) | 0.181 |  |  |
| Polluting cooking fuel | 1.20 (1.07 to 1.34) | 0.002 | 0.91 (0.82 to 1.02) | 0.101 | 1.09 (0.99 to 1.20) | 0.075 |  |  |
| Tube well (vs piped) | 1.08 (0.97 to 1.20) | 0.171 | 0.83 (0.76 to 0.91) | <0.001 | 1.06 (0.97 to 1.16) | 0.201 |  |  |
| Other water source (vs piped) | 1.06 (0.90 to 1.25) | 0.474 | 1.06 (0.80 to 1.40) | 0.702 | 1.28 (1.10 to 1.50) | 0.002 |  |  |
| Source water arsenic 10 to 49 ppb (vs below 10) |  |  |  |  |  |  | 1.47 (1.13 to 1.91) | 0.004 |
| Source water arsenic 50 ppb or more |  |  |  |  |  |  | 3.17 (2.44 to 4.11) | <0.001 |

Survey-weighted linear models of the log concentration, adjusted for all factors shown. The water test subsample model (n = 201) adjusts for sociodemographic factors and division only. P is the Wald test for that term.

### S6 Table. Blood metals, haemoglobin and anaemia (arsenic, cadmium and mercury terms exploratory)

| Model | Exposure | Estimate (95% CI) | P |
| --- | --- | --- | --- |
| Haemoglobin, g/dL difference | Log blood lead | -0.01 (-0.12 to 0.10) | 0.849 |
| Haemoglobin, g/dL difference | Blood lead 3.5 µg/dL or more | 0.07 (-0.15 to 0.29) | 0.516 |
| Anaemia, prevalence ratio | Blood lead 3.5 µg/dL or more | 0.93 (0.80 to 1.09) | 0.396 |
| Anaemia, prevalence ratio | Log blood lead | 1.02 (0.94 to 1.11) | 0.624 |
| Haemoglobin, four metals together | Log lead | 0.01 (-0.10 to 0.12) | 0.885 |
| Haemoglobin, four metals together | Log arsenic | -0.18 (-0.29 to -0.06) | 0.002 |
| Haemoglobin, four metals together | Log cadmium | -0.09 (-0.20 to 0.02) | 0.110 |
| Haemoglobin, four metals together | Log mercury | 0.14 (0.01 to 0.27) | 0.028 |

Anaemia (WHO 2024 trimester-specific cut-offs) was 53.6% (50.7 to 56.5) in women below 3.5 µg/dL and 51.0% (44.0 to 58.0) in women at or above it. All models adjust for age, education, parity, trimester, wealth, area and division.

### S7 Table. Blood lead in pregnant women and a child aged 12 to 59 months in the same household

| Estimate | All pairs (n = 540) | P | Woman is the child’s mother (n = 454) | P |
| --- | --- | --- | --- | --- |
| Spearman correlation | 0.37 | <0.001 | 0.38 | <0.001 |
| Geometric mean, woman (µg/dL) | 1.98 (1.87 to 2.11) |  | 2.04 (1.91 to 2.18) |  |
| Geometric mean, child (µg/dL) | 4.57 (4.31 to 4.84) |  | 4.58 (4.30 to 4.87) |  |
| Child to woman ratio (geometric mean) | 2.30 (2.15 to 2.47) |  | 2.24 (2.08 to 2.42) |  |
| Change in log child lead per log woman lead, crude | 0.29 (0.19 to 0.40) | <0.001 | 0.30 (0.19 to 0.42) | <0.001 |
| Change in log child lead per log woman lead, adjusted | 0.24 (0.13 to 0.35) | <0.001 | 0.25 (0.14 to 0.37) | <0.001 |
| Child at 5 µg/dL or more if woman below 3.5, % | 39.3 (33.9 to 44.9) |  | 39.2 (33.4 to 45.4) |  |
| Child at 5 µg/dL or more if woman at 3.5 or more, % | 67.5 (53.7 to 78.8) |  | 67.7 (53.6 to 79.2) |  |
| Adjusted prevalence ratio for child at 5 µg/dL or more | 1.50 (1.16 to 1.96) | 0.002 | 1.51 (1.15 to 1.98) | 0.003 |
| Adjusted slope, also adjusted for household soil lead (n = 537) | 0.23 (0.12 to 0.33) | <0.001 |  |  |
| Soil lead slope in the same model | 0.13 (-0.01 to 0.27) | 0.077 |  |  |

Adjusted models include the child’s age, wealth, area and division. Children were higher than the woman in their household in 486 of 540 pairs. P is blank where no formal test applies (a geometric mean or a ratio of means).

### S8 Table. Blood lead in pregnant women by district (city corporations merged into their districts)

| District | Women tested, n | Lead ≥3.5 µg/dL, % (95% CI) | Lead ≥5 µg/dL, % | Geometric mean, µg/dL |
| --- | --- | --- | --- | --- |

District estimates rest on 10 to 60 women each and are imprecise; the wide intervals show this. They are for description only.

### S9 Table. Blood cadmium and mercury in pregnant women (exploratory)

| Metal | Geometric mean (95% CI) | Median | 95th percentile | Maximum | At lowest reported value, n | Benchmark | % (95% CI) |
| --- | --- | --- | --- | --- | --- | --- | --- |
| Cadmium (µg/L) | 0.63 (0.61 to 0.65) | 0.65 | 1.78 | 34.1 | 8 | ≥1 µg/L (descriptive) | 24.5 (22.4 to 26.8) |
| Mercury (µg/L) | 1.45 (1.40 to 1.51) | 1.41 | 4.31 | 21.7 | 27 | ≥5.8 µg/L (methylmercury benchmark) | 2.2 (1.6 to 3.1) |

The survey report withheld these metals because of concerns about detection limits and precision. No assay or quality control data were available. Mercury is probably total blood mercury, so the methylmercury benchmark is only a concentration reference.

### S10 Table. Probability of a valid blood test among eligible pregnant women, and weight diagnostics

| Factor | Odds ratio (95% CI) | P |
| --- | --- | --- |
| Age 20 to 24 | 0.93 (0.68 to 1.26) | 0.631 |
| Age 25 to 29 | 0.92 (0.63 to 1.36) | 0.683 |
| Age 30 to 49 | 0.99 (0.64 to 1.52) | 0.948 |
| Secondary education | 0.80 (0.58 to 1.08) | 0.144 |
| Higher secondary or above | 0.51 (0.36 to 0.73) | <0.001 |
| One previous birth | 1.40 (1.05 to 1.87) | 0.022 |
| Two or more previous births | 1.22 (0.84 to 1.78) | 0.296 |
| Second trimester | 1.00 (0.78 to 1.29) | 0.990 |
| Third trimester | 0.66 (0.51 to 0.87) | 0.002 |
| Wealth: second | 0.72 (0.51 to 1.03) | 0.075 |
| Wealth: middle | 0.68 (0.47 to 0.97) | 0.034 |
| Wealth: fourth | 0.66 (0.46 to 0.95) | 0.025 |
| Wealth: richest | 0.38 (0.26 to 0.56) | <0.001 |
| Urban | 1.10 (0.86 to 1.39) | 0.457 |
| Chattogram | 0.31 (0.19 to 0.49) | <0.001 |
| Dhaka | 0.25 (0.16 to 0.40) | <0.001 |
| Khulna | 0.31 (0.19 to 0.52) | <0.001 |
| Mymensingh | 0.32 (0.17 to 0.60) | <0.001 |
| Rajshahi | 0.46 (0.27 to 0.79) | 0.005 |
| Rangpur | 0.80 (0.46 to 1.38) | 0.423 |
| Sylhet | 0.42 (0.24 to 0.73) | 0.002 |
| Hindu household | 0.98 (0.65 to 1.48) | 0.928 |

Logistic model weighted by the women’s design weight (n = 2,709 with complete covariates). Reference groups: age 15 to 19, primary or no education, no previous birth, first trimester, poorest quintile, rural, Barishal. Predicted probability of a valid test ranged from 0.33 to 0.95 (median 0.76). Response weights relative to their mean ranged from 0.06 to 5.25 (1st and 99th percentiles 0.11 and 3.48). The survey blood weight ranged from 0.05 to 3.98 times its mean. In our check, the blood weight equalled the women’s weight times the inverse of the stratum response rate, rescaled.

### S11 Table. Composition of eligible and tested women, and estimates with response weights

| Characteristic | All eligible, women’s weight (%) | Tested, blood weight (%) | Tested, response weight (%) |
| --- | --- | --- | --- |
| **Age** |  |  |  |
| 15 to 19 | 25.1 | 25.0 | 25.3 |
| 20 to 24 | 31.7 | 31.0 | 31.8 |
| 25 to 29 | 23.8 | 23.4 | 23.5 |
| 30 to 49 | 19.4 | 20.6 | 19.4 |
| **Education** |  |  |  |
| Primary or none | 18.6 | 20.9 | 18.7 |
| Secondary | 54.8 | 56.6 | 54.9 |
| Higher secondary or above | 26.6 | 22.5 | 26.5 |
| **Wealth quintile** |  |  |  |
| Poorest | 18.1 | 20.1 | 18.1 |
| Second | 19.9 | 20.5 | 19.8 |
| Middle | 20.1 | 20.3 | 20.1 |
| Fourth | 22.5 | 22.7 | 22.5 |
| Richest | 19.5 | 16.4 | 19.5 |
| **Area** |  |  |  |
| Urban | 29.5 | 29.3 | 29.7 |
| Rural | 70.5 | 70.7 | 70.3 |
| **Division** |  |  |  |
| Barishal | 5.6 | 5.7 | 5.6 |
| Chattogram | 25.9 | 26.2 | 25.9 |
| Dhaka | 25.0 | 24.8 | 25.1 |
| Khulna | 9.8 | 9.6 | 9.7 |
| Mymensingh | 6.5 | 6.5 | 6.5 |
| Rajshahi | 10.7 | 10.6 | 10.7 |
| Rangpur | 9.8 | 9.9 | 9.9 |
| Sylhet | 6.7 | 6.7 | 6.7 |
| **Trimester** |  |  |  |
| First | 30.3 | 30.6 | 30.4 |
| Second | 41.4 | 42.7 | 41.4 |
| Third | 28.3 | 26.7 | 28.2 |

| Estimate | Survey blood weight | Response weight | Response weight, trimmed |
| --- | --- | --- | --- |
| Blood lead ≥3.5 µg/dL, % | 17.7 (15.7 to 19.9) | 17.7 (15.8 to 19.8) | 17.6 (15.7 to 19.6) |
| Blood lead ≥5 µg/dL, % | 7.6 (6.1 to 9.3) | 7.6 (6.3 to 9.2) | 7.6 (6.3 to 9.2) |
| Geometric mean, µg/dL | 2.05 (1.99 to 2.12) | 2.05 (1.99 to 2.12) | 2.05 (1.98 to 2.11) |
| Greater Dhaka, ≥3.5 µg/dL, % | 65.2 (55.4 to 73.9) | 63.9 (53.9 to 72.8) | 64.4 (54.6 to 73.2) |
| Share of women at ≥3.5 living in greater Dhaka, % | 44.6 (38.1 to 51.2) | 48.1 (42.4 to 53.9) | 47.9 (42.2 to 53.6) |
| Share of women at ≥5 living in greater Dhaka, % | 63.6 (53.5 to 72.7) | 67.0 (57.9 to 74.9) | 66.8 (57.7 to 74.8) |
| Model 2: rest of Dhaka division vs greater Dhaka, PR | 0.30 (0.21 to 0.44) | 0.28 (0.20 to 0.41) | 0.28 (0.20 to 0.40) |
| Model 2: other divisions vs greater Dhaka, PR | 0.16 (0.12 to 0.22) | 0.16 (0.11 to 0.21) | 0.15 (0.11 to 0.21) |
| Model 2: richest vs poorest quintile, PR | 1.29 (0.82 to 2.05) | 1.23 (0.76 to 1.99) | 1.22 (0.76 to 1.96) |
| Pairs: child ≥5 if woman ≥3.5, adjusted PR | 1.50 (1.16 to 1.96) | 1.52 (1.16 to 1.99) | 1.54 (1.18 to 2.01) |

Response weights divide the women’s design weight by the modelled probability of a valid test (S10 Table). Trimmed weights are capped at the 1st and 99th percentiles. Model 1 estimates with response weights are in S4 Table.

### S12 Table. Greater Dhaka by component district

| Area (MICS district code) | Women tested, n | Clusters, n | Lead ≥3.5, n | Lead ≥5, n | Lead ≥3.5, weighted % (95% CI) |
| --- | --- | --- | --- | --- | --- |
| Dhaka South City Corporation (20) | 18 | 15 | 16 | 11 | 87.1 (57.4 to 97.1) |
| Dhaka North City Corporation (25) | 23 | 14 | 14 | 9 | 63.7 (39.6 to 82.4) |
| Dhaka (26) | 18 | 15 | 14 | 9 | 75.9 (49.2 to 91.1) |
| Gazipur (33) | 28 | 20 | 12 | 7 | 45.8 (29.3 to 63.3) |
| Narayanganj (67) | 25 | 21 | 19 | 11 | 69.8 (46.7 to 85.9) |
| **Greater Dhaka** | 112 | 85 | 75 | 47 | 65.2 (55.4 to 73.9) |

Share of all tested pregnant women in greater Dhaka: 12.1 (10.5 to 13.9)%. Share of women at ≥3.5 µg/dL: 44.6 (38.1 to 51.2)%. Share of women at ≥5 µg/dL: 63.6 (53.5 to 72.7)%. The grouping was defined after inspecting district estimates and is exploratory. Counts are unweighted; percentages are survey weighted.

### S13 Table. Pregnant women with and without a tested child in the household

| Characteristic | With a tested child (n = 540) | Without (n = 1,392) |
| --- | --- | --- |
| Geometric mean blood lead, µg/dL | 1.98 (1.87 to 2.11) | 2.08 (2.00 to 2.16) |
| Blood lead ≥3.5 µg/dL, % | 14.6 (11.2 to 18.7) | 18.8 (16.4 to 21.5) |
| Urban, % | 26.3 (22.1 to 31.0) | 30.4 (27.9 to 33.1) |
| Greater Dhaka, % | 7.3 (4.7 to 11.2) | 13.8 (11.8 to 16.1) |
| Fourth or richest wealth quintile, % | 36.0 (31.0 to 41.4) | 40.2 (37.1 to 43.3) |
| At least one previous birth, % | 90.7 (87.4 to 93.2) | 52.1 (48.9 to 55.2) |
| Mean age, years | 24.7 (24.2 to 25.2) | 24.2 (23.8 to 24.6) |

Raised lead in paired versus unpaired women, adjusted for place and wealth: PR 0.96 (0.73 to 1.25), P=0.742. With the child’s blood weight, the adjusted PR for a child at ≥5 µg/dL when the woman was at ≥3.5 µg/dL was 1.58 (1.22 to 2.04) (P<0.001). The adjusted log-log slope was 0.25 (0.14 to 0.36). Pair results apply to households with a pregnant woman and a tested child aged 12 to 59 months.

### S14 Table. Model size, overlap among site items and a simpler model

Model 1 has 31 terms for 254 women with blood lead ≥3.5 µg/dL. The largest variance inflation factor in a linear model with the same terms was 3.06 (mean 1.79). For the site items it ranged from 1.11 to 2.00.

| Item pair | Correlation (phi) |
| --- | --- |
| Battery and Jewellery | 0.45 |
| Battery and E-waste | 0.40 |
| Battery and Auto repair | 0.45 |
| Battery and Contact works at a site | 0.12 |
| Jewellery and E-waste | 0.45 |
| Jewellery and Auto repair | 0.64 |
| Jewellery and Contact works at a site | 0.26 |
| E-waste and Auto repair | 0.46 |
| E-waste and Contact works at a site | 0.15 |
| Auto repair and Contact works at a site | 0.20 |

Simpler model for blood lead ≥3.5 µg/dL (place, wealth, education, age and trimester; n = 1,921)

| Factor | PR (95% CI) | P |
| --- | --- | --- |
| Rest of Dhaka division (vs greater Dhaka) | 0.28 (0.20 to 0.40) | <0.001 |
| Other divisions (vs greater Dhaka) | 0.15 (0.12 to 0.21) | <0.001 |
| Wealth: second | 1.35 (0.86 to 2.13) | 0.190 |
| Wealth: middle | 1.29 (0.84 to 1.98) | 0.247 |
| Wealth: fourth | 1.14 (0.74 to 1.76) | 0.557 |
| Wealth: richest | 1.31 (0.84 to 2.05) | 0.238 |
| Secondary education | 1.00 (0.76 to 1.32) | 0.988 |
| Higher secondary or above | 0.73 (0.52 to 1.04) | 0.084 |
| Age 20 to 24 | 1.07 (0.78 to 1.48) | 0.658 |
| Age 25 to 29 | 1.45 (1.03 to 2.04) | 0.034 |
| Age 30 to 49 | 1.22 (0.85 to 1.73) | 0.277 |
| Second trimester | 1.03 (0.79 to 1.33) | 0.830 |
| Third trimester | 1.03 (0.77 to 1.38) | 0.850 |

Survey-weighted Poisson model with robust variance. Reference groups as in Table 3.

### S15 Table. Geometric means with and without values at the lowest reported level

| Metal | Values at lowest level, n | Geometric mean, all | Geometric mean, excluding these values (95% CI) |
| --- | --- | --- | --- |
| Lead (µg/dL) | 6 | 2.05 | 2.07 (2.01 to 2.14) |
| Arsenic (µg/L) | 2 | 2.12 | 2.12 (2.04 to 2.21) |
| Cadmium (µg/L) | 8 | 0.63 | 0.64 (0.62 to 0.66) |
| Mercury (µg/L) | 27 | 1.45 | 1.49 (1.44 to 1.54) |

Values at the lowest reported level were used as reported, without substitution. Cut-off based prevalences are unaffected because these values lie below every cut-off.
